# Remote monitoring of physical activity captured by the MyHeart Counts app identifies pulmonary arterial hypertension

**DOI:** 10.1101/2025.04.08.25325466

**Authors:** Juan A Delgado-SanMartin, Niamh Errington, Narayan Schuetz, Anders Johnson, Daniel Seung Kim, Varsha Gupta, Steve Hershman, Mark Toshner, Martin R Wilkins, David G Kiely, Roger Thompson, Euan Ashley, Dennis Wang, Allan Lawrie

**Affiliations:** National Heart and Lung Institute, Imperial College London, London, UK; Department of Medicine, Stanford University, CA, USA; Agency for Science, Technology and Research (A*STAR), Singapore; University of Texas, Austin, USA; Department of Medicine, University of Cambridge, UK; National I/HPAH Cohort Study, UK; NIHR Biomedical Research Centre, Sheffield and University of Sheffield, Sheffield, UK

**Keywords:** real-world data, remote monitoring, personal activity, machine learning, pulmonary hypertension

## Abstract

**Introduction:** The diagnosis of Pulmonary arterial hypertension (PAH) is often delayed. Wearable devices and smartphone applications have potential to remotely monitor patients and as access to technology increases, potentially identify patients at risk of cardiovascular disease, and reduce diagnostic delays.

**Methods:** We utilised data from 109 UK participants with up to 8 years of real-world activity monitoring captured using the MyHeart Counts app. We analysed physical activity, heart rate data and questionnaire responses from patients with idiopathic PAH (IPAH, n=34), disease controls (n=14), and healthy individuals (n=61). Validation studies were performed in an age and sex matched US MyHeart Counts cohort (28 IPAH, 23 other cardiovascular disease controls, and 22 healthy individuals).

**Results:** There were significant differences in activity levels between patients with PAH and controls. Activity metrics improved for patients with PAH following diagnosis. Correlations were observed between wearable-derived metrics and clinical variables such as 6MWT distance and NT-proBNP levels, supporting their potential to complement traditional risk assessment tools. Utilising pre-diagnostic data, we developed a classifier to identify patients with PAH from controls (ROC AUC = 0.87±0.07). The addition of patient responses to questionnaires, particularly on lifestyle enhanced model performance (ROC AUC = 0.94±0.07). The model was then tested in a US MyHeart Counts validation cohort. Following re-training (accounting for differences in control population) the model provided a performance of ROC AUC = 0.74±0.11.

**Conclusion:** This study highlights the feasibility of using wearable and smartphone technology to enable early detection, longitudinal monitoring, and remote risk assessment in PAH.

## INTRODUCTION

Pulmonary hypertension (PH) represents a significant yet often overlooked cardiovascular condition characterised by elevated mean pulmonary artery pressure (mPAP) greater than 20 mmHg (1). Pulmonary arterial hypertension (PAH), a rare form of PH, can develop in isolation (idiopathic or heritable), or in association with other diseases e.g. systemic sclerosis, congenital heart disease. In all instances PH imparts a poor prognosis and regardless of cause is commonly underdiagnosed. As with most rare diseases (2) there is often a significant delay (2-3 years) from first symptom to diagnosis, and subsequent treatment, especially since early symptoms are similar to those of other heart and lung conditions (3). We have previously demonstrated that this delay can result in patients presenting with more advanced disease, often complicated by the accumulation of other co-morbidities (particularly in older patients), resulting in presentation at high-risk of 1-year mortality (4). Reducing the time to diagnosis is therefore important to improve patient outcome and reduce the number of investigations performed, and allow treatment at an earlier stage of disease where therapies may be more effective (3). There are several clinical screening and early detection algorithms in use for patients at risk of PAH e.g. systemic sclerosis (3) but screening for idiopathic forms of PAH (IPAH) remains a challenge.

Under the current care model, patient information is only collected during infrequent clinic visits, and even frequent visits can be mistimed and ineffective for tracking disease progression (5). Smartphone applications (apps) and wearable technology offers the opportunity to remotely monitor and collect information on patients longitudinally outside of the clinic, such as their home (6). A range of cardiovascular diseases have shown a positive association between increasing activity levels and positive outcomes, including reduced mortality rates (7), and a lack of physical activity is a major risk factor in the development of heart disease. Despite the growing body of literature in this field and healthcare impact, there have been little effort to study the relationship between long-term real-world physical activity and PH using wearable devices (8). Studies using wearables with accelerometers have a short period of activity monitoring, for example, a fixed period of 5 days (9) or a week (10), and do not provide insight into everyday life or the patient’s journey before and after diagnosis. In PH, previous studies in controlled study conditions have reported a lower baseline level of physical activity in patients with PAH (9,11) and averaged step count has been found to associate with quality of life (QoL) in PH (11,12).

The MyHeart Counts Cardiovascular Health Study was launched in 2015 as one of the first studies to utilise the open source ResearchKit framework from Apple Inc (Cupertino, CA, USA) to facilitate clinical research entirely via a smartphone. The MyHeart Counts app (MHC) is free to download. It incorporates e-consent, HealthKit measurements of physical activity and fitness, as well as questionnaire assessment of sleep, lifestyle factors, risk perception, mental well-being, overall well-being and a 6-minute walk test (6MWT) (13). MHC has been utilised for studies in the US (13–16) as well as the UK (17) for real-world longitudinal studies for cardiovascular and COVID-19 related studies.

We present a study utilising passively collected real-world activity data and questionnaire responses in a cohort of PAH patients, disease controls, and healthy volunteers. Data from 109 participants spanning 273 days to 8.4 years was analysed. We provide an analysis of longitudinal wearable signals (heart rate, VO2Max, physical activity, sleep, etc.) in relation to PAH symptoms and questionnaire responses related to perception and mindset about physical activity and lifestyle. These data identify potential early warning signals from smartphones and wearables that can be correlated to traditional clinical variables and used to develop a PAH classification model. The classification model was validated in an external cohort of patients in the US to propose putative digital biomarkers for longitudinal, remote patient monitoring in PAH.

## METHODOLOGY

### UK Participant Recruitment

Following our IRB-approved protocol, 157 participants were recruited from either the Sheffield Teaching Hospitals Observational Study of Pulmonary Hypertension, Cardiovascular and other Respiratory Disease (STH-ObS) (UK REC Ref 18/YH/0441), or the UK National Cohort of idiopathic and heritable PAH study (H/IPAH Cohort) (NCT019072950) following informed written consent across four specialist PH centres in the UK (Royal Hallamshire Hospital (Sheffield), Hammersmith Hospital (London), Royal Papworth Hospital (Cambridge), and Royal United Hospital (Bath)). Eligible participants (over the age of 18 years) in possession of an Apple iPhone 6 or later were provided with a study pseudo-ID, offered an Apple Watch Series 4, invited to download the MyHeart Counts iOS app from the Apple App Store https://apps.apple.com/us/app/myheart-counts/id972189947, and provide additional consent into the MyHeart Counts study (NCT03090321). Baseline demographics, medical history, and relevant clinical parameters were obtained from STH-ObS or H/IPAH clinical databases. The demographics of UK participants is shown in Table 1. Participants were asked to wear/carry their devices consistently, and if possible, to wear their Apple Watch overnight. MyHeart Counts data is securely transmitted to a centralised Synapse database. Upon consenting into the MyHeart Counts study, historical HealthKit data stored on device e.g. step count is retrospectively collected.

**Table 1:** UK MHC participant demographics.

| UK |  |  |  |  |
| --- | --- | --- | --- | --- |
|  | PH | Healthy | Disease Control | Total |
| Gender |  |  |  |  |
| Female | 26 | 41 | 3 | 70 |
| Male | 8 | 20 | 11 | 39 |
| Ethnicity |  |  |  |  |
| White | 25 | 51 | 10 | 86 |
| Non-white | 3 | 3 | 1 | 7 |
| Unknown | 6 | 7 | 3 | 16 |

### US Validation Cohort

73 participants from the US MHC Stanford Cohort were used as an external validation cohort, of which 28 self-reported to have PAH, 23 self-reported other cardiovascular diseases (used as diseased controls) and 22 self-reported to be healthy, see their complete demographics in *Table 2* and the distribution of data compared to the UK cohort can be seen in the Supplementary Figure SA1.

**Table 2:**
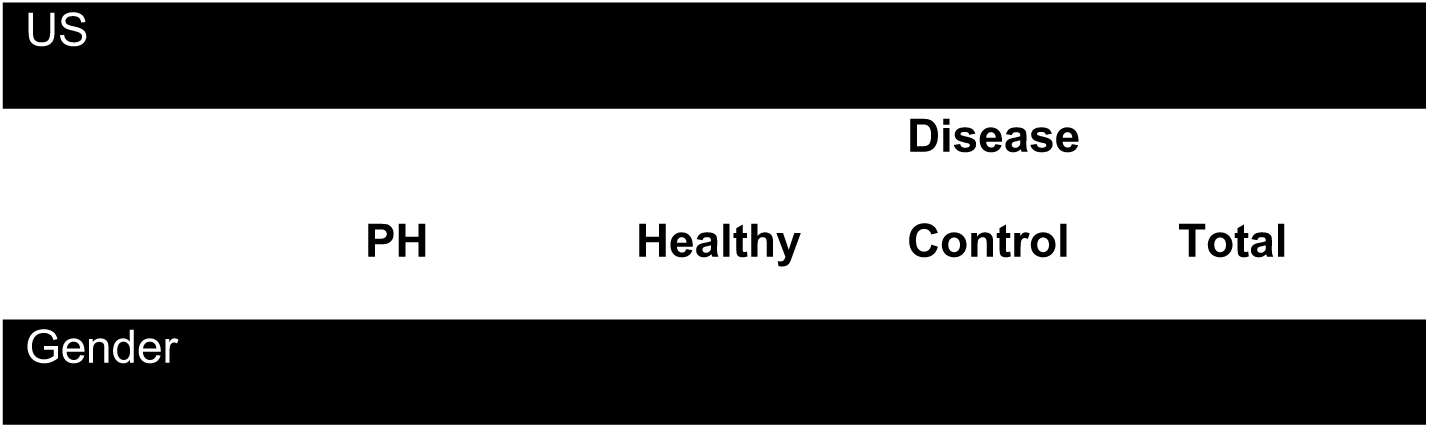
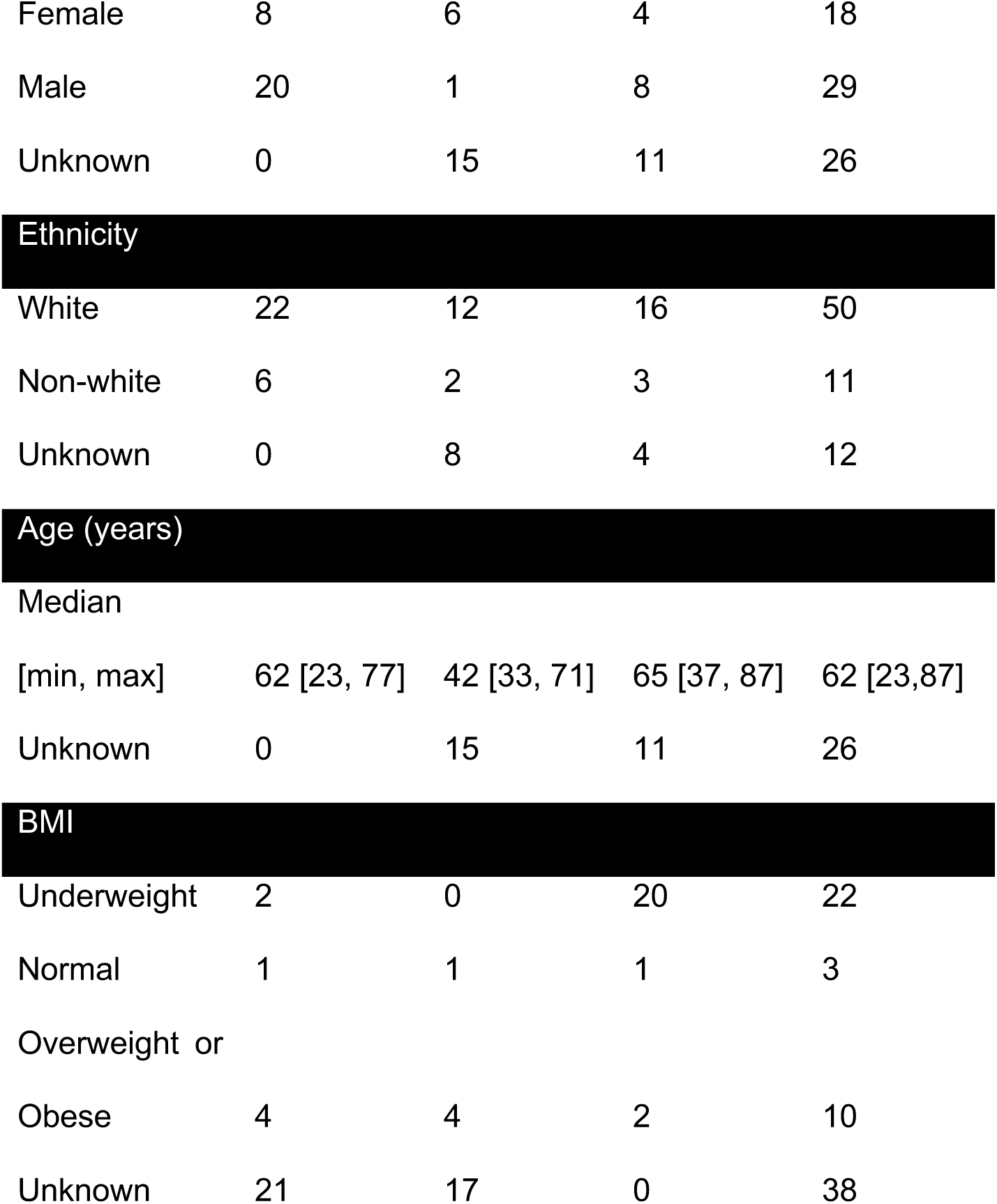
US MHC participant demographics (external validation). Data was obtained from HealthKit which had missing values.

### Data processing

MyHeart Counts Data was parsed with each user’s PseudoID to enable matching to their clinical data. HealthKit data were processed and cleaned to correct for duplicates, outliers & missing data prior to aggregated at hourly and daily levels. Duplicates arising from software updates or queues in software pipelines (about 15% of the data) were removed. Outliers were regarded as values too low or too large to be achieved in a single day were removed (about 13.7% of the data, see Table SA1). Errors in formatting of dates, or not-a-number values accounted for less than 0.01% of the data and missing data from non-wear or non-use was not included in this computation. For complete details on data processing refer to appendix A2. Details on questionnaire data can be found in Appendix D1. Aggregated data were then linked to PH clinical phenotype data, questionnaire responses and uploaded to Google Cloud using a BigQuery data warehouse. For the machine learning classifier, we ran a featurization pipeline which includes macro-statistical features, frequency-domain stats, and ARIMA features to all variables at a monthly aggregation level by patient.

### Physical activity variables measured by iPhone and Apple Watch

Descriptive statistics are used to summarise the variable characteristics of the study population. Variables included: Time Awake (hrs), Time In Bed (hrs), stepCount (steps), Time Asleep (hrs), appleStandTime (hrs), flightsClimbed, distanceWalkingRunning (m) present in both iPhone and Apple Watch; vo2Max (*mL ⋅ min/Kg*), heartRateVariabilitySDNN (ms), Average daily heartrate (beats/min), restingHeartRate (beats/min), walkingHeartRate Average (daily, beats/min), basalEnergyBurned (KCal), activeEnergyBurned (KCal) present on Apple Watch; and derived metrics (note derived metrics are capitalised): HeartRateReserve (beats/min), CardiacEffort (beats/m whilst walking), StepCountPaceMax or StepCountPaceMean (maximum/average daily gait speed), FlightsClimbedPaceMax or FlightsClimbedPaceMean (maximum/average daily speed climbing stairs), calculated as follows:

### Heart Rate Reserve

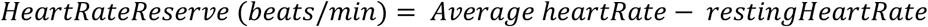

### Cardiac Effort

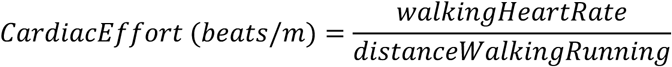

### Walking/climbing Pace (Gait speed)

First, duration was calculated by EndTime – StartTime in seconds. Any duration of less than 0.5 seconds was excluded. For any count of steps or flights climbed, we divided the steps by duration:

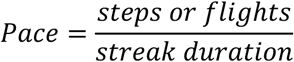

The sleep data presented here is first-party only, we did not use any third-party app or devices to monitor sleep. The algorithm used to process the data and new computed metrics is provided in supplemental material appendix A2. For the assessment of distribution similarity, we used a U-Mann-Whitney frequentist test and linear mixed effects model including all demographic information, time as fixed covariates, and the Group as random covariates, i.e. ‘value ∼ age + bmi + gender + ethnicity + months + prediagnosis’. The slopes of trajectories have been calculated in 3-month intervals, 12 months either side of diagnosis per patient. A one-way ANOVA test has been run to assess differences between intervals in the slopes.

### Self-reported lifestyle and mindset variables

In the first 7-days following enrolment into the MHC study participants are requested to complete a number of surveys including Physical Readiness Questionnaire (PAR-Q), Activity and Sleep survey (18), Cardio Diet Survey, Well-being and Risk perception Survey, as previously described (18), and exercise and activity process mindset (19). The questionnaires can be completed multiple times, but few participants did. We therefore focused analysis on the data captured in the first 7-days and treated as a static dataset. Kruskal-Wallis tests were applied to variables with Likert scales to identify significant differences between the 3 categories of participants (Pulmonary Arterial Hypertension (PH) disease controls (DC), and healthy controls (HC)) and P values adjusted using FDR correction. Fisher’s exact test was applied for variables with binary responses, using simulated p values in R (4.3.1). Post-hoc Dunn tests were carried out for question responses with a significant (<0.05) p value (dunn.test v1.3.6).

Polychoric correlation matrices were calculated using the polycor package in R (v0.8-1) for each of the 3 mindset questionnaires (Illness mindset, exercise process mindset, and activity process mindset). Patients with >50% missing values across a mindset questionnaire were removed prior to calculation.

To deal with missing values prior to factor analysis of mixed data (FAMD), samples with > 50% missing values were removed. Missing values were then imputed using the imputeFAMD function from the missMDA (v1.19). The best number of components to use in the imputation process was estimated as 2 (utilising the estim_ncpFAMD function from the missMDA package). FAMD was then carried out using the FactoMineR package (v 2.11).

### Developing a classifier for PAH

#### Model training

To generate predictive models, we compared two Machine learning algorithms: Binary classification using XGBoost (xgboost==2.0.3) and Linear SVM models (scikit-learn==1.5.0). We performed an internal 5-fold cross-validation using UK data only, leaving a 30% hold-out for validation. Hyperparameters were chosen by Bayesian Optimization. The values used for XGBoost were: reg alpha :2, reg lambda :5, scale pos weight: 2^(−1)1-*i*^, learning rate: 0.07, max depth: 6, n estimators: 30.

#### Model selection

We selected the model based on weakest cross-validation i.e. the most conservative to compared receiver operating characteristic (ROC) curves, F1, precision and recall as evaluation metrics. The thresholds were chosen based on optimised ROC AUCs and F1 scores at a 0.1 granularity.

#### Model evaluation

We tested on two holdout sets: intrinsic (UK data) and extrinsic (US data, transfer learning). For the extrinsic validation, we retrained the models on all UK data using the US as test (see Figure SC1). Thresholds have been chosen to be maximal based on ROC AUCs and F1. Finally, we re-trained the models using a small subset of the US data (20%) to improve efficiency of transfer learning, making sure to exclude the 20% segment from the test set avoiding data leakage. We use the external validation cohort to characterise data drift in a real situation much like it should be done in ML systems design. We implemented Population Stability Index (PSI) to track any potential performance degradation. Finally, features importance was defined by Decision Tree Importance.

### Code availability

The code is available here: https://github.com/jadsm/myheartcounts_ph and the runtime

requirements were of at least 8GB RAM & 2.4GHz, although it is recommended to have higher specs. Parallelization is recommended. The work was developed on an Apple Silicon M3 96GB running python 3.11.7 using the virtual environment in Appendix E. Visualisations are interactive and have been done in Altair (altair==5.3.0) and deployed in Google App Engine (available at https://mhc-imperial.silico.science).

### Data availability

Data can be requested from corresponding author for research purposes after ethics and governance approval.

## RESULTS

### UK Participant recruitment and data availability

From the 157 participants recruited, 109 had HealthKit activity data collected over a 10-year period (2014-09-23 - 2024-08-29) and were included in our analysis. Participants with infrequent or no data available were excluded. 34 participants were diagnosed with IPAH (PH) with 1 patient withdrawing consent. 14 participants were considered as disease controls (DC) comprising 12 post-hospitalisations for severe COVID-19, and 2 patients with suspected PH but normal pulmonary artery pressure (<20 mmHg). The remaining 61 participants were considered healthy controls. From the 33 PH patients, 21 had data prior to their diagnosis (April 2015 to November 2022), whereas all 33 patients having data post-diagnosis, with ranges of from 6 days to 7 years between Aug 2016 and Aug 2024 (Figure 1 A&B). In addition to the activity data, 62 of those participants performed at least one on-app 6-minute walking test (MHC-specific 6MWT – not the estimated 6MWT), 57 participants provided sleep data, and 29 participants had recorded Apple Watch Workout data (not used) (Figure 1C). All 109 patients responded to at least 5 questions within the questionnaires to assess existing levels of physical activity, sleep, diet, health and mindset, with the majority answering most of them (Figure 1D). Supplementary Figure SD1 shows a breakdown of distribution of answers to questionnaires and Table SD1 the counts.

**Figure 1:**
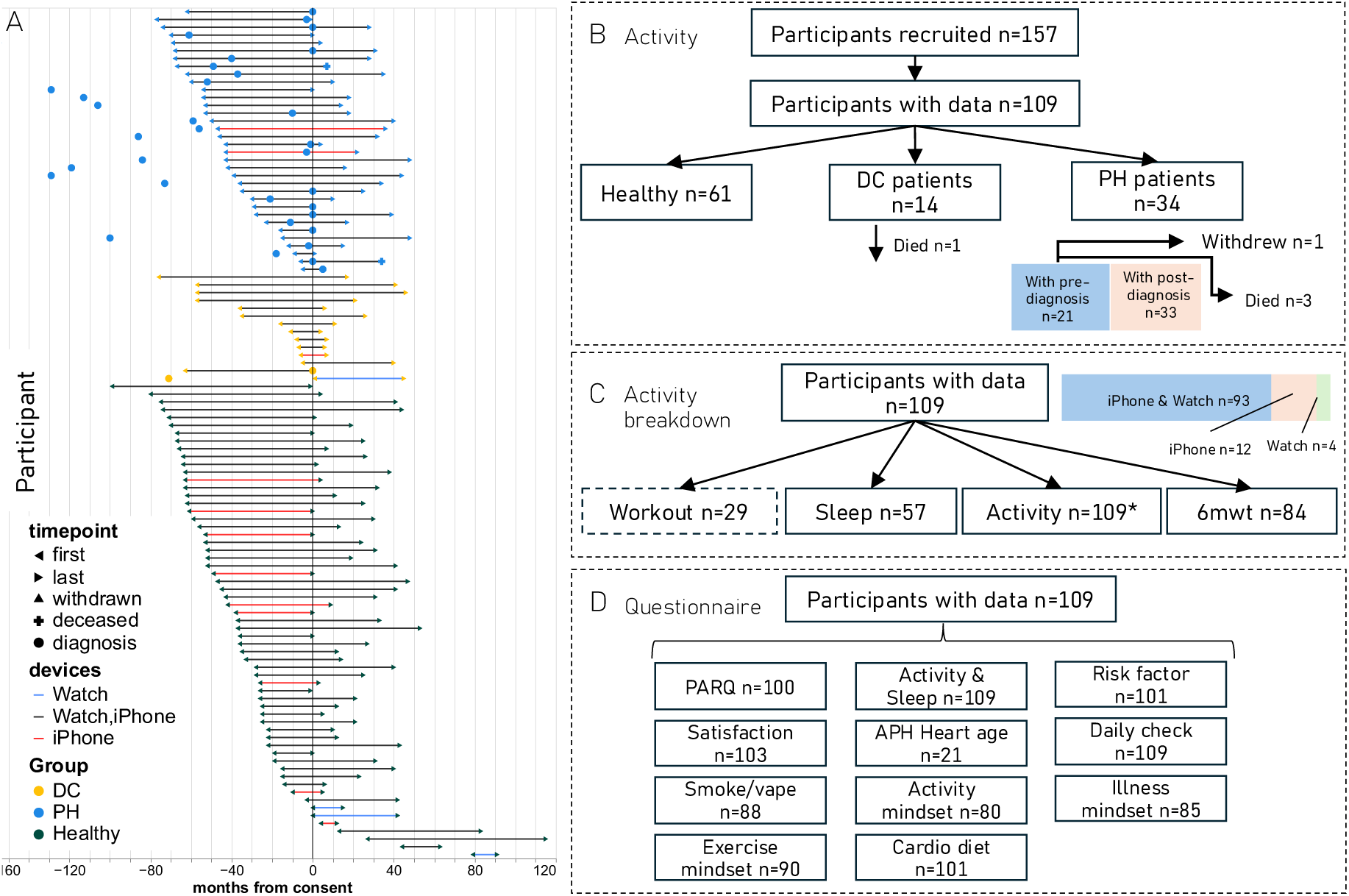
UK MyHeart Counts recruitment and data availability. A) Timeline of activity data in relation to date from consent into MyHeart Counts (0 months) and relative time of diagnosis. Participants are grouped into PH, DC or Healthy volunteers. The solid lines indicate where data were available within a 6-month sliding window, with the colour of the lines indicating the device(s) data were obtained from. B) Summary of the participants with activity data within each group. C) Breakdown of activity sources. D) number of participants who provided input to each questionnaire. Dotted line represents data not used in our analyses; DC, disease controls; PH, idiopathic pulmonary arterial hypertension; 6mwt, six-minute walking test; PARQ, physical activity readiness questionnaire.

### UK MHC Cohort Clinical data

Clinical records for all 33 PH patients were linked to the MHC data via the provided pseudoID. Basic demographic data were available from clinical databases and reported in Table 1. PH phenotype characteristics were obtained from those participants that underwent a diagnostic right heart catheterisation for diagnosis of PH and are summarised in Table 3.

**Table 3:** Clinical Phenotype data for UK MHC participants at time of diagnosis given as Mean ± Standard Deviation.

| Right Heart Catheter |  |  |
| --- | --- | --- |
| mPAP (mmHg) | Mean Pulmonary Artery Pressure | 53.6 ± 15.6 |
| PAWP (mmHg) | Pulmonary Wedge Pressure | 8.4 ± 3.5 |
| PVR (Dynes.sec.cm<sup>-5</sup>) | Pulmonary Vascular Resistance | 1042.2 ± 569.2 |
| Cardiac Output (l/min) | | 3.8 ± 1.8 |
| Lung Function |  |  |
| FEV1% Predicted | Forced Effective Volume 1 | 85.8 ± 15.2 |
| FVC% Predicted | Forced Vital Capacity | 93.4 ± 18.3 |
| TCLO% Predicted | | 62.6 ± 18.4 |
| Exercise Test |  |  |
| 6MWD (m) | 6-minute walk test | 450.9 ± 114.9 |
| ISWD (m) | Intermittent Shuttle test | 514.4 ± 275.1 |
| BNP/NT-proBNP |  |  |
| NT-proBNP (pg/ml) | N-terminal pro B-Type<br>Natriuretic Peptide | 471.2 ± 664.4 |
| BNP (ng/L) | B-Type Natriuretic Peptide | 195.2 ± 217.4 |
| WHO FC I/II/III/IV (%) | World Health<br>Organisation's Functional<br>Class | 6/30/63/1 |

### Phone accelerometer-derived HealthKit activity metrics reveal differences between participant groups and identifies longitudinal trends pre-and post-diagnosis

We first chose to investigate the utility of the activity data captured solely via the accelerometer on the iPhone. The distribution of physical activity measures from HealthKit across all time points for each participant identified significant differences across the participant groups for stepCount (p-value<0.001) and flightsClimbed (p-value <0.001) where patients with IPAH demonstrated a reduced number of both metrics (Figure 2, left column). Patients with IPAH also climbed less flights of stairs and walked less than the healthy and DC groups. From these two HealthKit features we calculated the average and maximum walking (StepCountPaceMean, StepCountPaceMax), and flight (stairs) climbing pace (FlightsClimbedPaceMean, FlightsClimbedPaceMax) (Figure 2, right column). The maximum pace was calculated to estimate the maximum effort achievable by each participant. Again, patients with IPAH walked and climbed flights slower than healthy individuals, with a maximum daily difference between healthy and PH of 32 ±10 steps/min and 0.6 ±0.6 flights/min.

**Figure 2:**
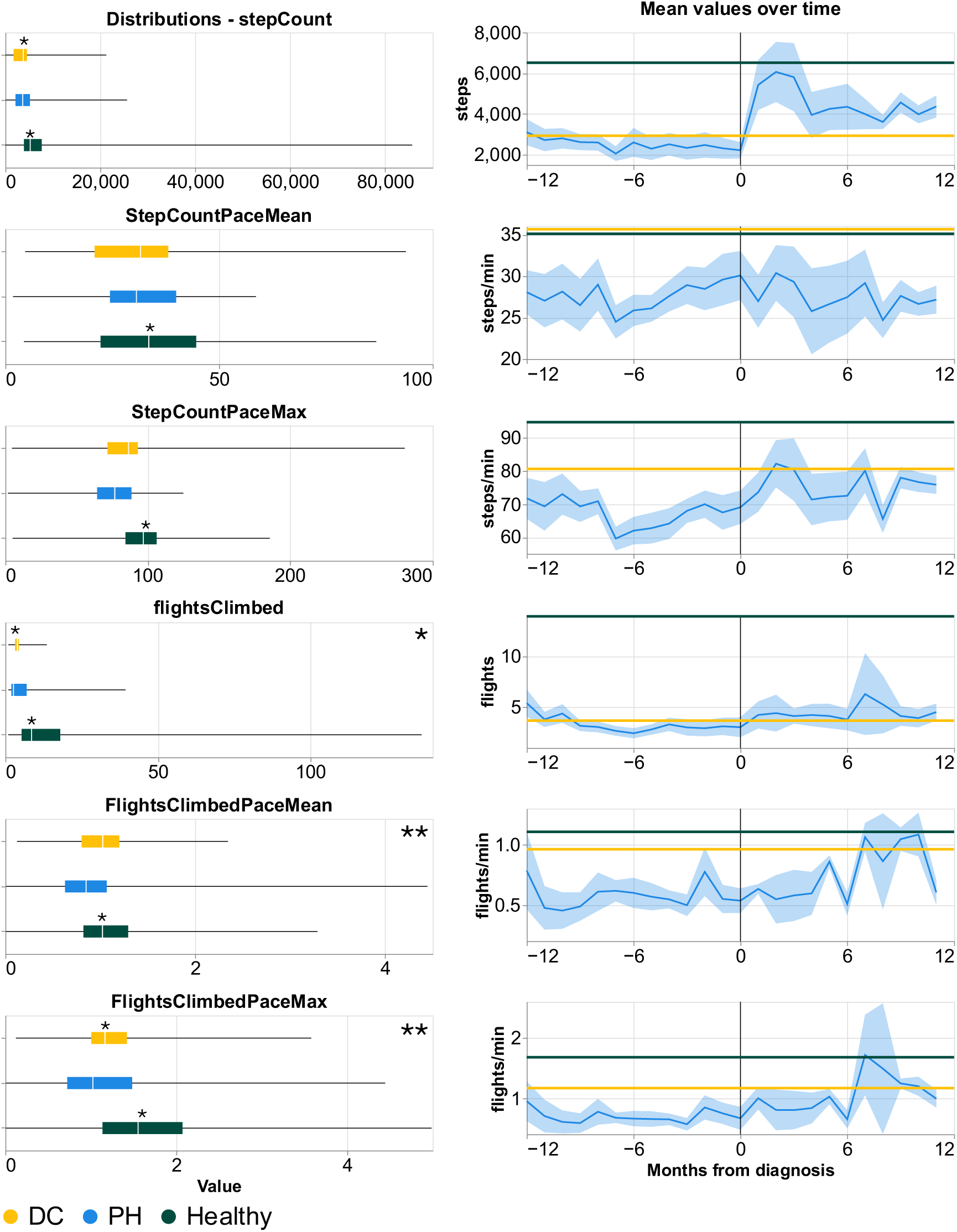
Distribution of physical activity variables measured in iPhone HealthKit. Distributions of HealthKit values prior to diagnosis in PH case only with top 0.5% removed (left column), and monthly mean values with 95% CI 12 months before and after diagnosis (right column). Lines for DC and Healthy represent the average of the group in the whole period. *U-Mann-Whitney significance P<0.05.

We next examined activity patterns to determine whether they were different pre- and post-diagnosis of IPAH. We saw that patients with IPAH improved after diagnosis across all metrics (Supplemental Figure SB1), particularly stepCount, AvgWalkPace, FlightsClimbed, AvgFlightPace and MaxFlightPace approaching averages similar to the disease control and healthy individuals (Figure 2 right column) with p-values of <0.05 (see supplementary Table SA2). Analysis of data collected within the first six months post-diagnosis highlighted a significant improvement in walking speed. We also observed a period of response immediately after diagnosis up to around 6 months (see Figure SB1).

### Differences in activity measures from smartwatch before and after PAH diagnosis

Since participants were offered an Apple Watch at the point of recruitment (most commonly diagnosis), and only a few participants already owned a device, there was less pre-diagnostic watch-based HealthKit features than pre-diagnostic phone derived accelerometer data. We first explored the watch-based accelerometer metrics (those shared with iPhones). Similar to the phone derived data we observed a pronounced separation between PH and Healthy groups in StepCountPaceMean with a steady increase in flightsClimbed and FlightsClimbedPaceMean over time following diagnosis (Figure 3A). There was a more gradual increase in gait speed (StepCountPaceMean) that was slightly more pronounced immediately after diagnosis before slowing and eventually decreasing around 9-10 months post-diagnosis (Figure 3A). There was a steady increase in appleStandTime after diagnosis in patients with PAH.

**Figure 3:**
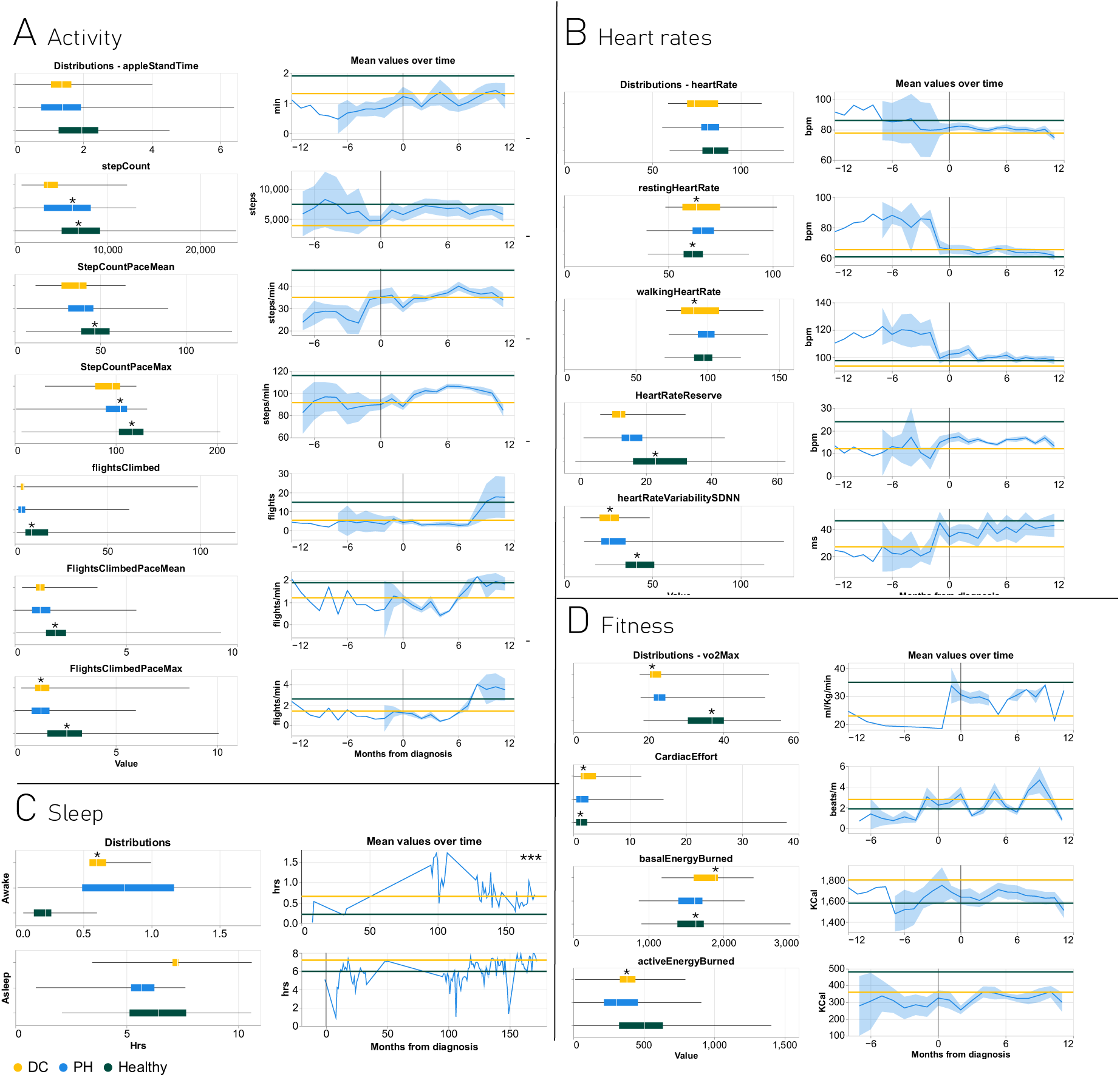
description of watch metrics: distributions of overall values with top 0.5% removed (left column), and monthly mean values with 95% CI 12 months before and after diagnosis (right column, lines for DC and Healthy represent the average of the group in the whole period. *Significance as measured by U-Mann-Whitney p<0.05. ***Non-linear mixed effect model shows effect. Variables are grouped in 4 categories: Activity (A), Heart rates (B), Sleep (C), and Fitness (D).

Examination of the data obtained from the PPG e.g. heart rates, identified significant differences in restingHeartRate, walkingHeartRateAverage and HeartRateReserve (Calculated from Average heartRate – restingHeartRate, Figure 3B) between patients with PAH and controls. There was a steep decline in both restingHeartRate and walkingHeartRate immediately prior diagnosis that was associated with an increase in HeartRateReserve. These changes were maintained following diagnosis. Data on sleep metrics was relatively sparse compared to other data. However, analysis of sleep data identified a significant difference in the time patients with PAH spent awake (56 min) at night compared to the control groups (15 min, Figure 3C). The Apple Watch estimates Basal and Active Energy burned in addition to VO_2_max based on the PPG sensor and activity. There was no significant difference between participant groups either before or after diagnosis (Figure 3D). We also calculated Cardiac Effort (average heart rate/time spend active) and identified a significant difference between patients with PH and the control groups, and variation within the patients with PH in the 12 months following diagnosis suggestive of cycles of improvement and decline in cardiovascular fitness (Figure 3D). We have also observed a period of response immediately after diagnosis which bounces back at around 6 months (see Figure SB2).

When looking at seasonality, we see a clear pattern of activity at commute and lunch time which is much more pronounced in healthy individuals (Figure SB3) with StepCountPaceMean of 31 (DC), 40 (Healthy), and 34 (PH) steps/min while commuting versus 28 (DC), 32 (Healthy) 30 (PAH) steps/min in the evening. When looking at times of the year, we see a clear pattern of activity increase in the warmer months which is tracked by all cohorts (Figure SB4), generating a starker difference for patients with PH compared to other groups with astepCount of 5460 (DC), 7760 (Healthy), and 5378 (PH) in winter versus 5772 (DC, +6%), 8087 (Healthy, +4%), and 6100 (PH, +13%) in spring. Finally, when looking at day of the week (Figure SB5), we see that healthy individuals on average increase their step count on the weekend but they move slower (average of 7855 steps at 47.9 steps/min on working days and 8092 steps at 45.3 steps/min on weekends), in contrast to PH patients who do not show significant differences between weekdays and weekends (average of 5834 steps at 38.9 steps/min on working days and 5449 steps at 38.2 steps/min on weekends).

### Differences in perception of physical activity and lifestyle factors between healthy and PH patients

Over the first 7-days following enrolment into the MHC study participants were requested to complete a number of surveys including Physical Readiness Questionnaire (PAR-Q), Activity and Sleep survey (18), Cardio Diet Survey, Well-being and Risk perception Survey (20), as previously described (13,16), and exercise mindset (19).

Statistical analysis (Kruskal-Wallis test) across all questionnaire responses to compare PH, Healthy and DC responders identified 15 questions (excluding questions directly related to cardiovascular disease and prescription drugs) with significant differences ranging from *pvalue* = 0.039 for ‘*chronic illness handling’* and whether they work or not (*pvalue* = 6.3 ⋅ 10^%^), see supplementary Table SD2. Of these variables the most prominent were related to risk factors and whether participants were physically capable of working. On the psychological mindset, the proportion of participants who found exercising relaxing, perception of their own bodies ability to heal, and that a chronic illness is a vehicle to find more meaning in life were significantly different across the groups (supplementary Table SD2). We subsequently performed a Factor Analysis of Mixed Data (FAMD) and found a significant separation in the distribution of responders that aligned with their participant group when projected onto a lower dimensional space (supplementary Figure SD2) suggesting that PH patients have different attitudes towards chronic diseases and have different lifestyle factors.

### Remote monitoring of physical activity, heart rate and questionnaire metrics correlate with clinical risk score of PAH progression

There are several clinical risk scores used by physicians to predict the 1-year survival for PAH (compared in Yogeswaran et al (21)), with specific algorithms favoured by region/country. However, the inclusion of a six-minute walk distance, NT-proBNP/BNP concentration and WHO functional class are included in the risk calculators, with some allowance for missing variables.

Clinical risk scores are beneficial when assessing patients’ disease severity at diagnosis and treatment response but are limited by the single snapshot of data collection. We therefore examined the correlation between watch and phone metrics and clinical scores (ERS/ESC 4-strata risk score (1)). As clinical tests can be performed at various timepoints, we aggregated results within 3 months of each other for comparison with our device metrics.

Our clinical datasets contained exercise data from two walk tests: Incremental Shuttle Walk Test (ISWT) and the 6-min Walk Test (6MWT). The MHC app records a remote 6MWT, but due to poor concordance with clinical tests (primarily conducted during the COVID-19 pandemic) and limited test repetitions, a direct correlation was not feasible in this study. The clinical 6MWT distance correlated well with flightsClimbed (*ρ* = 0.53), HeartRateReserve (*ρ* = 0.63) and heartRateVariabilitySDNN (HRV) (*ρ* = 0.60), see Figure 4. There was also a negative correlation with restingHeartRate (*ρ* = −0.36) and basalEnergyBurned (*ρ* = −0.53). The ISWT distance similarly had a strong positive correlation with flightsClimbed (*ρ* = 0.33), stepCount (*ρ* = −0.52), HeartRateReserve (*ρ* = 0.59) and heartRateVariabilitySDNN (*ρ* = 0.66), and a strong negative correlation with average heartRate (*ρ* = −0.72), restingHeartRate (*ρ* = −0.86) and walkingHeartRate (*ρ* = −0.85) see Figure 4. Average heartRate provided the strongest negative correlation (*ρ* = −0.36) with NT-proBNP/BNP levels (Figure 4). Meaningful correlations between WHO functional class and activity and heart rate metrics proved to be more challenging, although some questionnaire response data, did provide some modest correlation (Supplementary Figure B4).

**Figure 4:**
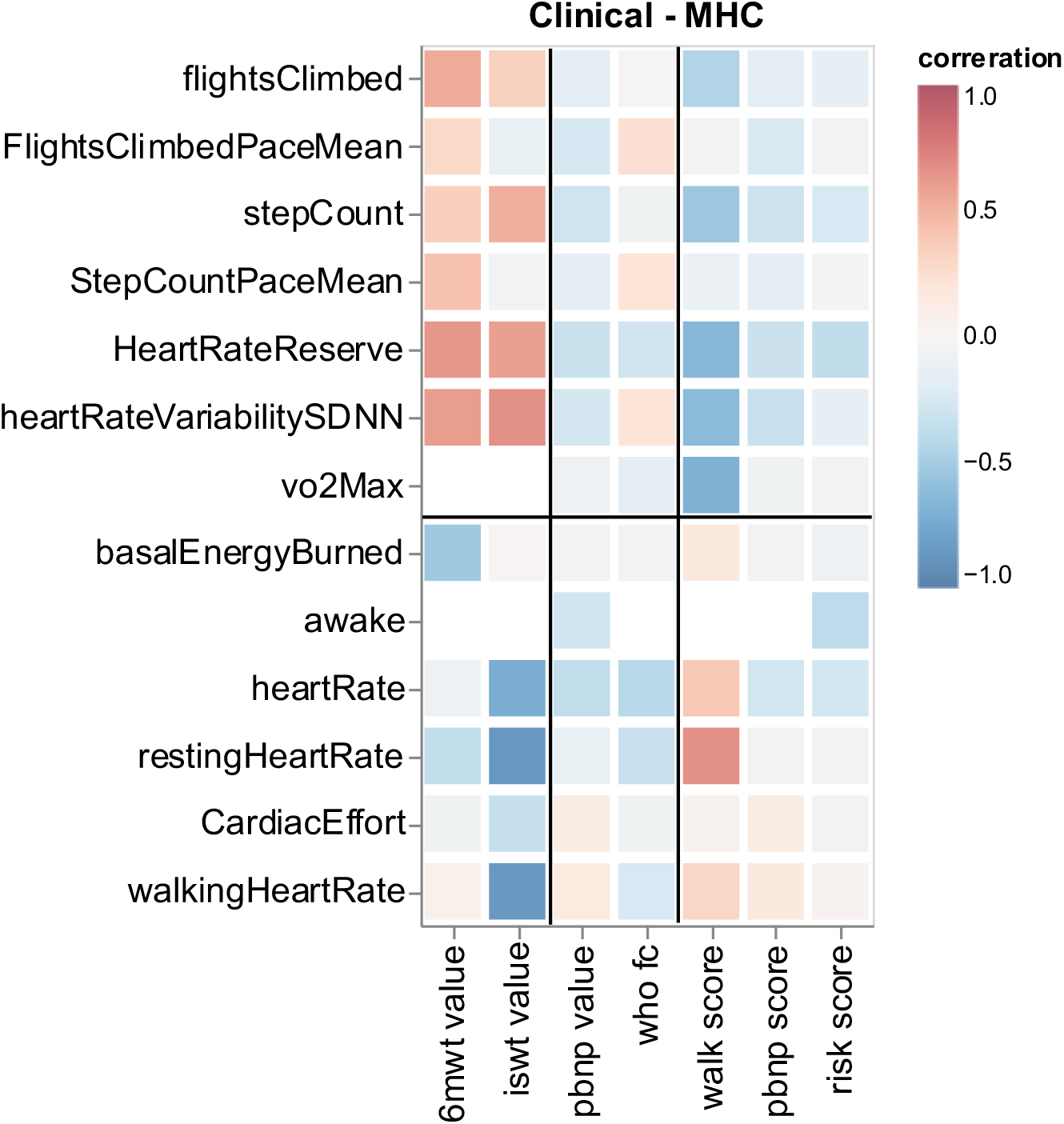
clinical risk assessment. Pearson’s correlation between clinical walk tests and risk scores with activity metrics pre-diagnosis. Labels in the x-axis represent 6-min walk test (6mwt), intermittent shuttle test (iswt), pro-BNP (pbnp), and WHO Functional Class (who fc).

We categorised the 6MWT distances and NT-proBNP/BNP levels based on thresholds from the ERS/ESC guidelines (1) and the ISWT distance using Lewis et al (22). After computing the walk score, we saw a strong positive correlation between higher risk walk scores and restingHeartRate, and strong negative correlations with vo2Max, and HeartRateReserve, stepCount and flightsClimbed (Figure 4). Average heartRate again provided the best negative correlation with NT-proBNP/BNP score (Figure 4). Despite significant correlations between the remotely collected activity, heart rate and questionnaire responses and individual constituents of the ERS/ESC 4-strata risk score, there was no significant correlation between the combined ERS/ESC risk score and any metric.

We computed correlations between activity-survey (Figure SB6 A) where we saw that awake, asleep, heartRateVariabilitySDNN, vo2Max, BMI, and height correlate with the most survey questions. We also looked at survey-clinical risk (Figure SB6 B), noting that the walk scores correlate best with questionnaire data. The surprising finding that the intermittent shuttle test and 6-minute walk test correlate with different variables underscores their distinct assessments of functional capacity. Notably, FlightsClimbedPaceMean and StepCountPaceMean correlate positively with 6-minute walk test (*ρ* = 0.28 and *ρ* = 0.42 respectively), but not with intermittent shuttle test (*ρ* = −0.13 and *ρ* = −0.04 respectively).

### Classifying PAH diagnosis using the patient’s physical activity and perception

Finally, we trained a binary classification model to classify patients with PAH from the combined disease and healthy controls. We trained two models using XGBoost, and linear SVM (comparisons in the Figure SC2). We separated the analysis into pre- and post-diagnosis to isolate diagnosis and treatment effect. Pre-diagnosis we achieve ROC AUCs of 0.87±0.08 (n=71) in the best case using all watch metrics, followed very closely by phone only data with ROC AUCs of 0.81±0.12 (n=92), see *Figure 5*A. In the post-diagnostic range the best ROC AUCs were 0.64±0.17 (n=96) for watch data and 0.48±0.18 (n=104) for phone data, 0.23 and 0.33 less than in the pre-diagnostic range (*Figure 5*B). This drop in performance matches expectations, since there are improvements in activity metrics in PAH patients following diagnosis.

**Figure 5:**
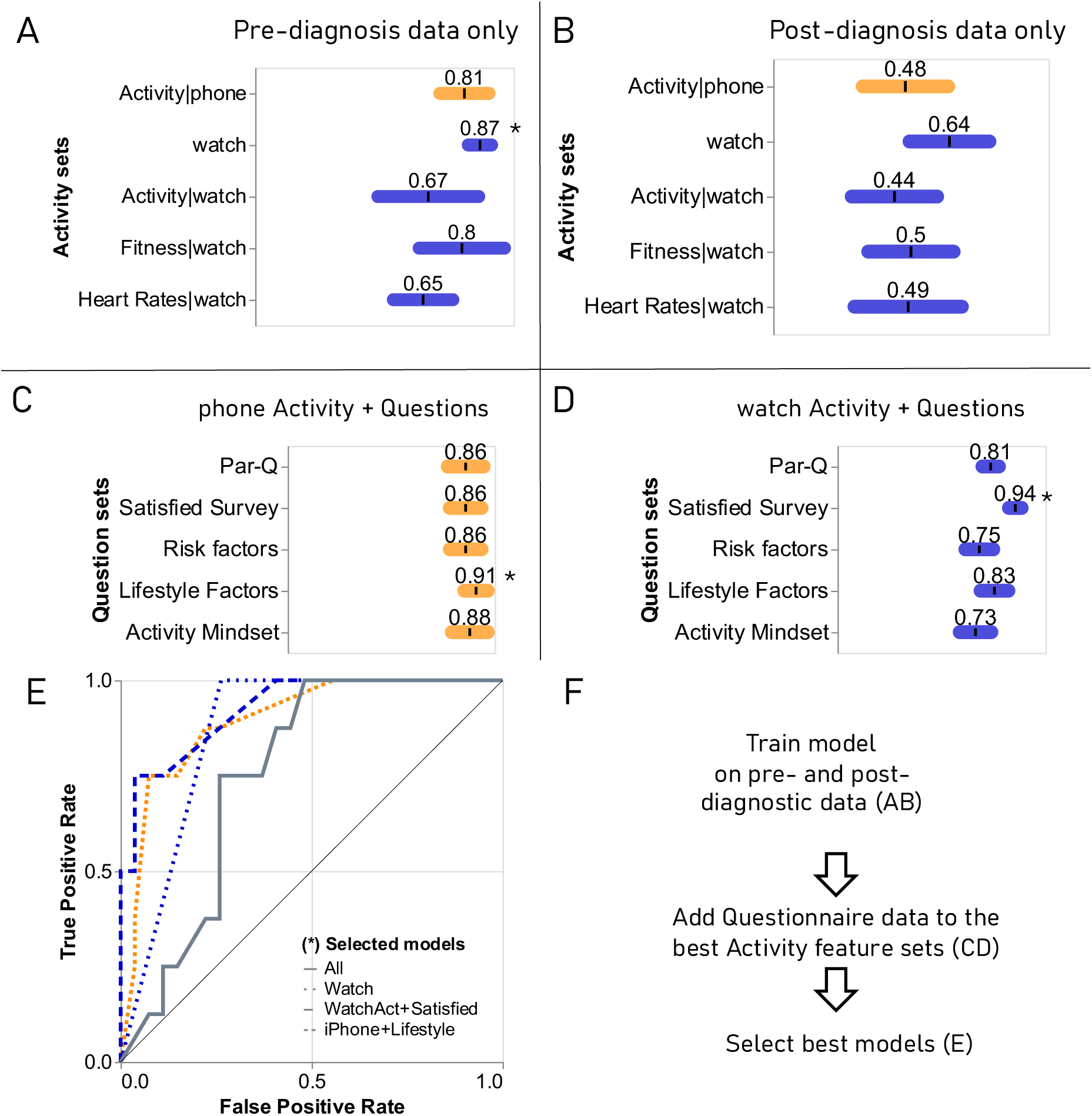
Model test ROC AUCs of intrinsic validation (UK cohort) for the best model XGBoost. A) ROC AUC mean ± stdev for activity data (pre-diagnosis). B) same results (post-diagnosis). *C)* pre-diagnostic results for the best activity models with sections of questionnaire for phone. *D)* Same pre-diagnostic results but for watch activity metric set. E) ROC curves of selected models (marked with *). F) Idea flow for the figure. Note: sleep data was insufficient to train a model (Table SC1).

We next focused on the pre-diagnostic range and added questionnaire features to the activity metrics for phone and watch based on 5 aggregations: PAR-Q (a standard physical activity readiness survey (23)), Satisfied survey (a psychological questionnaire on life and exercise satisfaction), Risk factors (risk factors to cardiovascular disease), Lifestyle factors (lifestyle factors such as diet and sleep), and Activity mindset (mindset towards physical activity), see Table and Figure SD3 for more details. The results show that phone AUCs improve uniformly with all questionnaires (see *Figure 5*C), whereas watch activity metrics see a variation in performance increase from 0.04 to 0.23 gains in ROC AUCs (see *Figure 5*D). From this, we conclude that the most promising models are the all the watch data, Activity watch+Satisfied Survey and phone+Lifestyle survey, all of which ROC have been illustrated in *Figure 5*E.

As judged by the decision tree importance, 61% of importance is attributed to flights climbed and step count metrics (see Table SC2).

### Testing the PAH classifier in an external US cohort

After training the model and testing on a UK hold-out data set (intrinsic validation), we decided to test the performance on a US cohort (extrinsic validation). Firstly, we observe that there is significant data drift in the activity and less pronounced in the questionnaire data (Figure SC3). When looking at individual activity variables we observed that the direction between groups is similar for UK and US, but the median and IQR values show some differences especially for healthy individuals (Figure SC4 for iPhone and SC5 for Apple Watch). For example, healthy individuals show StepCountPaceMax with medians and IQRs of 112 [100, 123] steps/min for UK and 86 [56,112] steps/min for US. As observed in Figure 6A, the model performed worse than random classifiers (ROC AUCs < 0.5). To overcome this data drift and generate a more generalised model, we included 20% of the US cohort into the training set of the model. After retraining, all models showed improved performance, reaching ROC AUCs of 0.74±0.12 for watch only, followed by 0.71±0.11 for Activity watch+Satisfied survey and 0.69±0.12 for phone+Lifestyle, demonstrating that even with only 20% of data (no further increase in performance was seen (Figure SC7)) from the new cohort, performance can be restored to levels comparable to the initial validation on Figure 5*A*.

**Figure 6:**
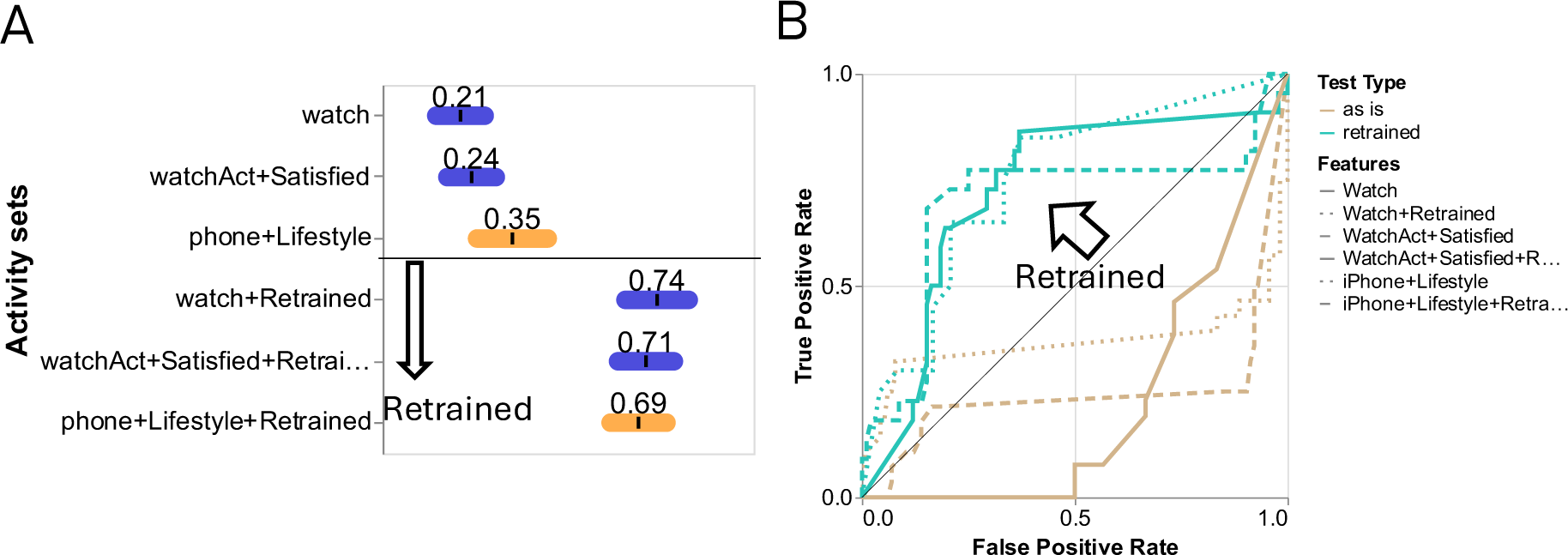
Model ROC AUCs for extrinsic validation (US cohort). A) ROC AUC mean ± stdev for each of the best models before and after retraining with 20% of US data. B) ROC curves for the same models.

## DISCUSSION

Our study highlights the potential of leveraging real-world activity and questionnaire data from smartphones and wearable devices to improve early detection and longitudinal monitoring of PAH. Our longitudinal data obtained via the MyHeart Counts iOS app demonstrates the utility of digital biomarkers in distinguishing patients with PAH from healthy and disease control groups, as well as tracking disease progression pre- and post-diagnosis.

We deliberately opted for a simplified framework, employing a basic feature extraction method (statistical and Fourier transform) and a relatively straightforward ML model (XGBoost). This conscious decision aimed to mitigate the challenges posed by the inherent noise encountered in real-world data. By minimizing the initial complexity, we sought to establish a robust baseline and gain a deeper understanding of the underlying data patterns before exploring more sophisticated techniques, such as longitudinal clustering methods like Latent Class Analysis or Growth Mixture Modelling, or more complex feature extractions like wavelet-based features. These data indicate that simple activity metrics derived from a smartphone such as step count, walking pace (gait speed), and pace at climbing flights provide valuable insights into physical activity patterns associated with PAH. Unsurprisingly, reduced activity levels and slower pace were evident in patients with PAH compared to healthy individuals, consistent with the functional limitations caused by the disease. Importantly, pre-diagnosis metrics, and the addition of wearable data showed promise for predicting PAH diagnosis, achieving an area under the curve (AUC) of 0.87±0.07 for watch-only model, and reaching 0.94+0.07 when adding Activity Satisfaction surveys. Although watch metrics resulted in better classification results, phone metrics showed very similar performance (AUC 0.91±0.09) but offers some advantages. For instance, the potential reach is much larger since most people (in 2025 about 90% of the world’s population is a smartphone user) already own a phone and have been using a phone for many years. This offers the possibility to retrospectively collect data years prior to diagnosis and the opportunity to understand disease related activity phenotypes). In the present work, the smartphone alone provided enough power and since more people have access to a phone over a watch, we can reach more people.

Our ML classifier was validated against an external population in the US. Importantly this contained participants with other cardiovascular diseases, and a healthy population with a different distribution of physical activity levels. After generalising the model by retraining the model with 20% of the US cohort, the model achieved acceptable performance for predicting PH diagnosis (AUC 0.74±0.11). These findings suggest that smartphone and wearable devices could play a critical role in identifying at-risk individuals and improving diagnostic delays for PAH.

Post-diagnosis, PAH patients demonstrated significant improvements in activity metrics, likely reflecting the benefits of treatment with treatment effects observed in other similar studies (24,25). Metrics such as walking pace and heart rate reserve approached levels obtained by healthy controls. Combining activity and heart rate metrics with patient-reported data further enhanced model performance, particularly for lifestyle and mindset-related surveys. This finding underscores the value of incorporating patient perspectives to contextualise activity data, offering a more comprehensive understanding of their health status. Further, psychological and lifestyle factors, including perceptions of physical capability and attitudes toward chronic illness, may provide additional diagnostic and prognostic insights.

While our study demonstrates the feasibility of using wearable and smartphone data for PAH monitoring, there are several limitations which we acknowledge. Firstly, the pre-diagnostic data capture was limited for certain metrics – particularly those related to overnight measurement, and there was variability in the duration and completeness of data across participants. Irregular gaps in data may reflect disease or individual specific habits, or even random patterns. In the design of our analysis, though, these gaps are unlikely to have a major effect, if we have a period of normal usage in the months immediately before and after diagnosis. However, gaps of over one month or multiple months can hinder the interpretation of our results. A more thorough analysis of individual patients can help answer these questions but requires a larger participant dataset. Moreover, there are significant challenges in comparing activity data collected in the UK and US populations. We suggest that there may be variability between UK and US populations related to lifestyle, which may include travel habits with US participants. The US participants also contained more non-PAH other cardiovascular disease patients, whereas the UK disease controls were largely post-hospitalised COVID patients, and thirdly the PH patients within the US population were self-reported. Despite these differences, we were able to demonstrate good transfer of knowledge to a completely different population including just 20% of data of the new population in the training. Finally, the observed decline patterns may not be specific to PH and could resemble those seen in other cardiovascular conditions, such as atrial fibrillation or obstructive coronary disease (26,27).

This manuscript demonstrates a foundational approach for the classification of PAH from a mixture of diseased controls and healthy participants. These non-invasive measures hold promise as a scalable, patient-centred solutions for empowering patients and doctors to track disease progression more closely. However, further research and validation in larger, more diverse cohorts are required before we can fully harness their potential and integrate them with other exposures that can impact upon health. These studies are critical if data from smartphone and wearables are to be incorporated into routine clinical care.

## Data Availability

All data produced in the present study are available upon reasonable request to the authors

## REFERENCES

1. Humbert M, Kovacs G, Hoeper MM et al. 2022 ESC/ERS Guidelines for the diagnosis and treatment of pulmonary hypertension. Eur Respir J 2023;61:2200879.

2. Phillips C, Parkinson A, Namsrai T et al. Time to diagnosis for a rare disease: managing medical uncertainty. A qualitative study. Orphanet Journal of Rare Diseases 2024;19.

3. Kiely DG, Lawrie A, Humbert M. Screening strategies for pulmonary arterial hypertension. Eur Heart J Suppl 2019;21:K9–K20.

4. Lawrie A, Hamilton N, Wood S et al. Association of risk assessment at diagnosis with healthcare resource utilization and health-related quality of life outcomes in pulmonary arterial hypertension. Pulmonary Circulation 2024;14.

5. Spatz ES, Ginsburg GS, Rumsfeld JS, Turakhia MP. Wearable Digital Health Technologies for Monitoring in Cardiovascular Medicine. N Engl J Med 2024;390:346–356.

6. Williams GJ, Al-Baraikan A, Rademakers FE et al. Wearable technology and the cardiovascular system: the future of patient assessment. Lancet Digit Health 2023;5:e467–e476.

7. Barbiellini Amidei C, Trevisan C, Dotto M et al. Association of physical activity trajectories with major cardiovascular diseases in elderly people. Heart 2022;108:360–366.

8. Lachant D, Light A, Lachant M et al. Peak steps: Capacity for activity improves after adding approved therapy in pulmonary arterial hypertension. Pulm Circ 2023;13:e12285.

9. Gonzalez-Saiz L, Santos-Lozano A, Fiuza-Luces C et al. Physical activity levels are low in patients with pulmonary hypertension. Ann Transl Med 2018;6:205.

10. Minhas J, Shou H, Hershman S et al. Physical Activity and Its Association with Traditional Outcome Measures in Pulmonary Arterial Hypertension. Ann Am Thorac Soc 2022;19:572–582.

11. Pugh ME, Buchowski MS, Robbins IM, Newman JH, Hemnes AR. Physical activity limitation as measured by accelerometry in pulmonary arterial hypertension. Chest 2012;142:1391–1398.

12. Lachant D, White RJ. Wearable Devices in Pulmonary Arterial Hypertension: What Are We Trying to Learn? Advances in Pulmonary Hypertension 2023;22:92–97.

13. McConnell MV, Shcherbina A, Pavlovic A et al. Feasibility of obtaining measures of lifestyle from a smartphone app: the MyHeart Counts Cardiovascular Health Study. JAMA cardiology 2017;2:67–76.

14. Javed A, Kim DS, Hershman SG et al. Personalized digital behaviour interventions increase short-term physical activity: a randomized control crossover trial substudy of the MyHeart Counts Cardiovascular Health Study. Eur Heart J Digit Health 2023;4:411–419.

15. Shcherbina A, Hershman SG, Lazzeroni L et al. The effect of digital physical activity interventions on daily step count: a randomised controlled crossover substudy of the MyHeart Counts Cardiovascular Health Study. The Lancet Digital Health 2019;1:e344–e352.

16. Hershman SG, Bot BM, Shcherbina A et al. Physical activity, sleep and cardiovascular health data for 50,000 individuals from the MyHeart Counts Study. Sci Data 2019;6:24.

17. Gupta V, Kariotis S, Rajab MD et al. Unsupervised machine learning to investigate trajectory patterns of COVID-19 symptoms and physical activity measured via the MyHeart Counts App and smart devices. NPJ Digit Med 2023;6:239.

18. Arena R, Arnett DK, Terry PE et al. The role of worksite health screening: a policy statement from the American Heart Association. Circulation 2014;130:719–34.

19. Boles DZ, DeSousa M, Turnwald BP et al. Can Exercising and Eating Healthy Be Fun and Indulgent Instead of Boring and Depriving? Targeting Mindsets About the Process of Engaging in Healthy Behaviors. Frontiers in Psychology 2021;12.

20. OECD. OECD Guidelines on Measuring Subjective Well-being OECD Publishing 2013.

21. Yogeswaran A, Gall H, Fünderich M et al. Comparison of Contemporary Risk Scores in All Groups of Pulmonary Hypertension. Chest 2024.

22. Lewis RA, Billings CG, Hurdman JA et al. Maximal Exercise Testing Using the Incremental Shuttle Walking Test Can Be Used to Risk-Stratify Patients with Pulmonary Arterial Hypertension. Ann Am Thorac Soc 2021;18:34–43.

23. Adams R. Revised Physical Activity Readiness Questionnaire. Can Fam Physician 1999;45:992, 995, 1004-5.

24. Sacks L, Kunkoski E. Digital Health Technology to Measure Drug Efficacy in Clinical Trials for Parkinson’s Disease: A Regulatory Perspective. J Parkinsons Dis 2021;11:S111–S115.

25. Elzinga WO, Prins S, Borghans L et al. Detection of Clenbuterol-Induced Changes in Heart Rate Using At-Home Recorded Smartwatch Data: Randomized Controlled Trial. JMIR Form Res 2021;5:e31890.

26. Somani S, Rogers AJ. Just in time: detecting cardiac arrest with smartwatch technology. Lancet Digit Health 2024;6:e148–e149.

27. Vyas R, Jain S, Thakre A et al. Smart watch applications in atrial fibrillation detection: Current state and future directions. J Cardiovasc Electrophysiol 2024;35:2474–2482.

