## Supplemental Material for "Remote monitoring of physical activity captured by the MyHeart Counts app identifies pulmonary arterial hypertension"

Supplementary materials of manuscript:

**The combination of remote monitoring of physical activity and its perception captured by the MyHeart Counts smartphone app identifies pulmonary arterial hypertension**

Juan A Delgado-SanMartin, Niamh Errington, Narayan Schuetz, Anders Johnson, Daniel Seung Kim, Varsha Gupta, Steve Hershman, Mark Toshner, Martin R Wilkins, David G Kiely, Roger Thompson, Euan Ashley, Dennis Wang, Allan Lawrie

### Appendix A: Details on Data and its analysis

##### A1 US cohort description

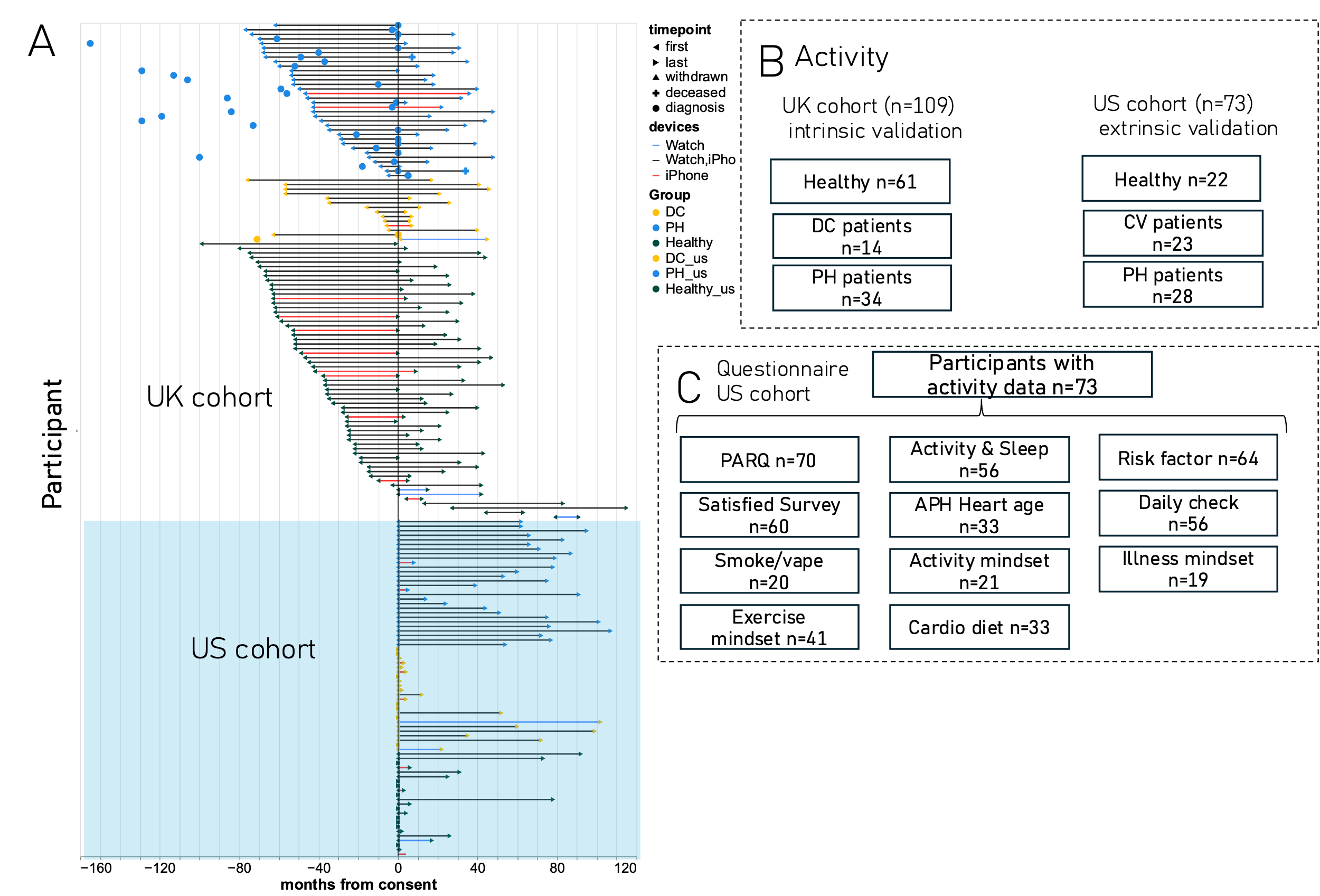

*Figure SA1: Description of US cohort in relation to UK cohort. A) timeline of participants. B) Participant counts for activity for UK and US cohorts. C) Questionnaire answer availability of US cohort with activity data.*

##### A2 Wearable Data Cleaning Pipeline

The cleaning pipeline consisted of the following operations:

1. Remove duplicate ids
2. Remove out-of-scope (oos) devices
3. Map devices
4. Remove oos variables
5. Aggregate
6. Device rank calculation
7. Remove out of bounds
8. Remove NaNs
9. Remove wrong dates
10. Calculate extra variables

Now in detail, one by one:

1. Duplicates have been removed by patient, startTime, endTime, type, and source.
2. Devices other than iPhone or Apple Watch were excluded
3. Devices were mapped from the HK ‘source’ using fuzzy logic.
4. The following variables were excluded:

InhalerUsage, BasalBodyTemperature, HighHeartRateEvent, MindfulSession, DistanceCycling, RespiratoryRate, OxygenSaturation, BodyTemperature, DietaryCholesterol, AppleStandHourIdle, AppleStandHourStood, BloodPressureDiastolic, BloodPressureSystolic

5. All variables were aggregated at an hourly and daily basis.
6. Device ranks were calculated on a daily basis – i.e. for each Apple ID, each device on any given day was given a rank. Later only top devices on any given day were included.
7. The bounds were set based on what is logical, all points outside of these bounds were considered outliers and excluded:

*Table SA1: bounds for each variable*

|  | **Lower bound** | **Upper bound** |  |
| --- | --- | --- | --- |
| **stepCount** | 50 | 100000 | steps |
| **flightsClimbed** | 0 | 500 | flights |
| **FlightsClimbedPaceMax** | 0 | 10000 | fligths / min |
| **FlightsClimbedPaceMean** | 0 | 10000 | fligths / min |
| **StepCountPaceMax** | 0 | 10000 | fligths / min |
| **StepCountPaceMean** | 0 | 10000 | fligths / min |
| **basalEnergyBurned** | 800 | 5000 | Kcal |
| **activeEnergyBurned** | 0 | 5000 | Kcal |
| **appleStandTime** | 0 | 24 | hours |
| **walkingHeartRateAverage** | 50 | 220 | beats / min |
| **heartRate** | 30 | 220 | beats / min |
| **restingHeartRate** | 30 | 220 | beats / min |
| **heartRateVariabilitySDNN** | 0 | 150 | ms |
| **vo2Max** | 0 | 60 | mL/Kg*min |
| **Height** | 1.4 | 2.2 | m |
| **BodyMass** | 40 | 200 | Kg |
| **distanceWalkingRunning** | 0 | 60 | m |
| **InBed** | 0 | 24 | Hours |
| **asleep** | 0 | 24 | Hours |
| **awake** | 0 | 24 | Hours |
| **CardiacEffort** | 0 | 100 | beats / m |

8. Not-a-number values were excluded.
9. Dates in the wrong format were excluded.
10. Extra variables were calculated as follows:
    - **Heart Rate Reserve**

$$HeartRateReserve= Average heartRate- restingHeartRate$$

- **Cardiac Effort**

$$CardiacEffort (beats/m)=\frac{walkingHeartRate}{distanceWalkingRunning}$$

- **Pace (Gait speed)**

First, duration was calculated by EndTime – StartTime in seconds. Any duration of less than 0.5 seconds was excluded. For any count of steps or flights climbed, we divided the steps by duration:

$$Pace=\frac{steps or flights}{duration}$$

- **BedBound**

Any number of hours above 18 hours of time in bed daily.

- **Time over effort threshold (70%)**

Time spent above 70% of their maximum effort.

The % drop of the data based on the last parsing (October 2024) is:

OOS devices: 0 rows - 0.0 %

OOS variables: 38323 rows - 1.7 %

Below lower bound: 65969 rows - 3.0 %

Above upper bound: 216359 rows - 10.2 %

Nans removed: 0 rows - 0.0 %

Illogical dates removed: 2 rows - 0.0 %

##### A3 Statistical analysis of wearable data

*Table SA2: p values of U-Mann-Whitney test for binary PH vs DC-Healthy with adjusted values*

|  |  |  | **pvalues adjusted** | |
| --- | --- | --- | --- | --- |
| **variable** | **device** | **groups** | **all** | **eth_age_gen** |
| flightsClimbed | iPhone | PH-DC | 3.5E-39 | 8.1E-99 |
| FlightsClimbedPaceMax | iPhone | PH-DC | 3.1E-02 | 3.0E-02 |
| FlightsClimbedPaceMean | iPhone | PH-DC | 4.8E-01 | 8.4E-01 |
| stepCount | iPhone | PH-DC | 8.1E-13 | 4.3E-06 |
| StepCountPaceMax | iPhone | PH-DC | 4.6E-01 | 4.4E-01 |
| StepCountPaceMean | iPhone | PH-DC | 5.6E-01 | 4.6E-01 |
| activeEnergyBurned | Apple Watch | PH-DC | 3.1E-09 | 5.5E-12 |
| appleStandTime | Apple Watch | PH-DC | 3.3E-01 | 1.1E-01 |
| asleep | Apple Watch | PH-DC | 1.0E-02 | 1.8E-02 |
| awake | Apple Watch | PH-DC | 5.6E-01 | 4.4E-01 |
| basalEnergyBurned | Apple Watch | PH-DC | 9.1E-16 | 1.5E-17 |
| CardiacEffort | Apple Watch | PH-DC | 3.3E-06 | 2.2E-04 |
| flightsClimbed | Apple Watch | PH-DC | 1.9E-02 | 4.0E-03 |
| FlightsClimbedPaceMax | Apple Watch | PH-DC | 6.5E-01 | 6.3E-01 |
| FlightsClimbedPaceMean | Apple Watch | PH-DC | 1.2E-02 | 6.1E-02 |
| heartRate | Apple Watch | PH-DC | 2.7E-01 | 7.9E-01 |
| HeartRateReserve | Apple Watch | PH-DC | 9.0E-02 | 2.4E-01 |
| heartRateVariabilitySDNN | Apple Watch | PH-DC | 3.6E-01 | 3.6E-09 |
| restingHeartRate | Apple Watch | PH-DC | 3.6E-06 | 1.2E-03 |
| stepCount | Apple Watch | PH-DC | 3.0E-05 | 1.2E-05 |
| StepCountPaceMax | Apple Watch | PH-DC | 1.0E-11 | 3.7E-11 |
| StepCountPaceMean | Apple Watch | PH-DC | 4.9E-07 | 1.3E-01 |
| vo2Max | Apple Watch | PH-DC | 2.9E-05 | 9.8E-08 |
| walkingHeartRateAverage | Apple Watch | PH-DC | 7.8E-04 | 1.5E-03 |
| activeEnergyBurned | iPhone | PH-Healthy | 5.3E-02 | 1.4E-01 |
| basalEnergyBurned | iPhone | PH-Healthy | 8.6E-05 | 1.4E-07 |
| flightsClimbed | iPhone | PH-Healthy | 1.6E-79 | 4.3E-28 |
| FlightsClimbedPaceMax | iPhone | PH-Healthy | 1.9E-03 | 8.2E-03 |
| FlightsClimbedPaceMean | iPhone | PH-Healthy | 1.4E-02 | 5.7E-03 |
| stepCount | iPhone | PH-Healthy | 5.8E-20 | 2.8E-19 |
| StepCountPaceMax | iPhone | PH-Healthy | 1.1E-02 | 1.1E-04 |
| StepCountPaceMean | iPhone | PH-Healthy | 4.5E-09 | 1.9E-08 |
| activeEnergyBurned | Apple Watch | PH-Healthy | 4.7E-01 | 1.5E-01 |
| appleStandTime | Apple Watch | PH-Healthy | 1.9E-01 | 2.2E-01 |
| asleep | Apple Watch | PH-Healthy | 5.5E-01 | 6.1E-01 |
| awake | Apple Watch | PH-Healthy | 1.4E-01 | 5.0E-02 |
| basalEnergyBurned | Apple Watch | PH-Healthy | 7.8E-15 | 1.7E-08 |
| CardiacEffort | Apple Watch | PH-Healthy | 2.4E-03 | 1.3E-02 |
| flightsClimbed | Apple Watch | PH-Healthy | 3.3E-01 | 1.8E-27 |
| FlightsClimbedPaceMax | Apple Watch | PH-Healthy | 6.6E-03 | 8.2E-02 |
| FlightsClimbedPaceMean | Apple Watch | PH-Healthy | 1.8E-02 | 1.0E-01 |
| heartRate | Apple Watch | PH-Healthy | 1.2E-03 | 3.0E-04 |
| HeartRateReserve | Apple Watch | PH-Healthy | 3.0E-09 | 2.3E-09 |
| heartRateVariabilitySDNN | Apple Watch | PH-Healthy | 5.8E-04 | 8.4E-14 |
| restingHeartRate | Apple Watch | PH-Healthy | 8.3E-01 | 7.4E-01 |
| stepCount | Apple Watch | PH-Healthy | 3.8E-02 | 7.4E-02 |
| StepCountPaceMax | Apple Watch | PH-Healthy | 1.8E-02 | 1.1E-08 |
| StepCountPaceMean | Apple Watch | PH-Healthy | 8.6E-01 | 3.6E-01 |
| vo2Max | Apple Watch | PH-Healthy | 2.8E-33 | 6.8E-33 |
| walkingHeartRateAverage | Apple Watch | PH-Healthy | 6.2E-02 | 6.9E-02 |

### Appendix B: Descriptive Meta-analysis

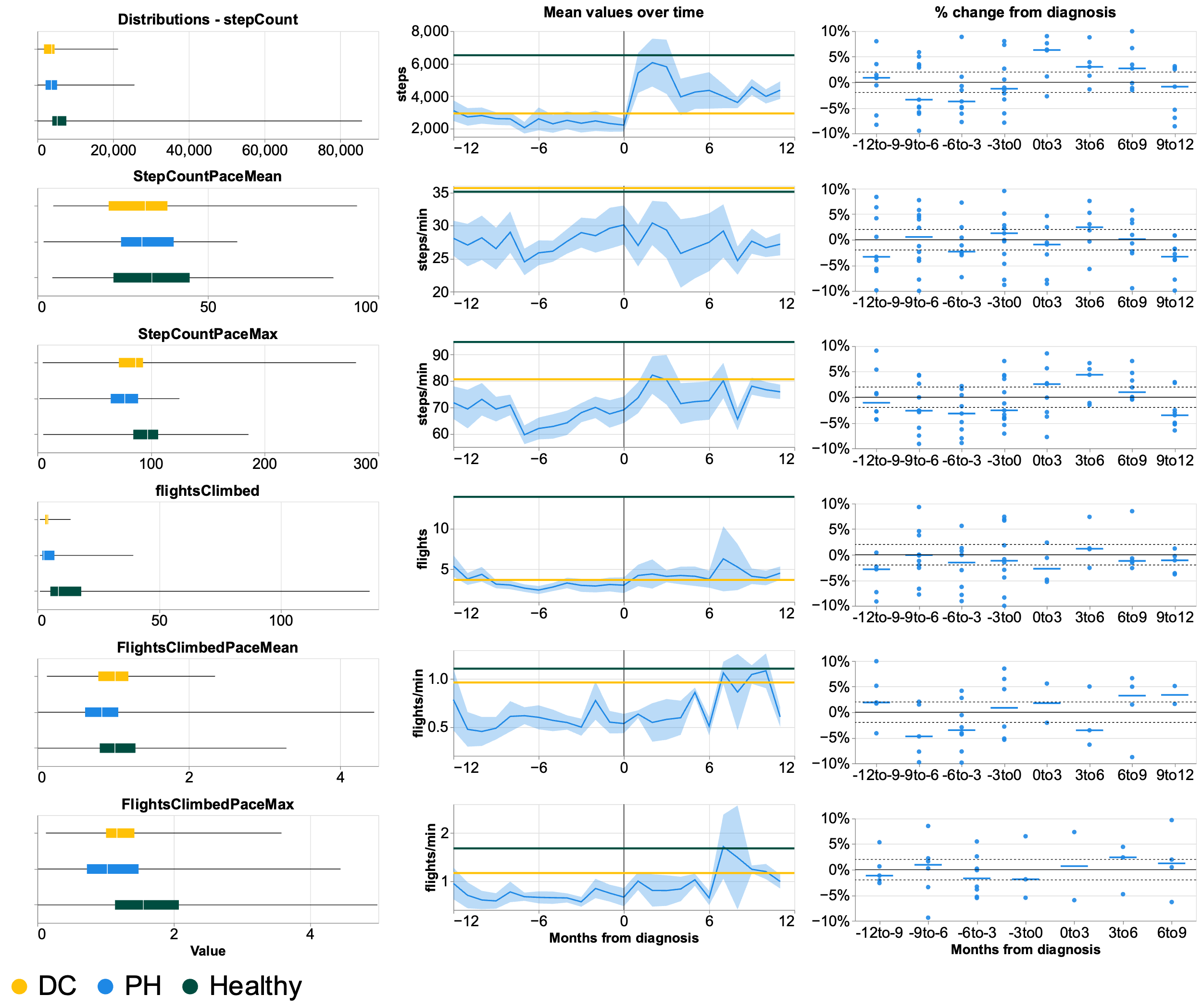

Figure SB1: description of iPhone metrics: distributions of values prior to diagnosis in PH case only with top 0.5% removed (left column), monthly mean values with 95% CI 12 months before and after diagnosis (right column, lines for DC and Healthy represent the average of the group in the whole period); and % change from diagnosis in 6 months intervals (right column, dots and strokes represent each patient and the median respectively). Non-linear mixed effect models show notable effect *, notable effect **. A one-way ANOVA test on the slopes between 12 and 6 month prior to diagnosis and 0-6 months after diagnosis shows 90% significance $.

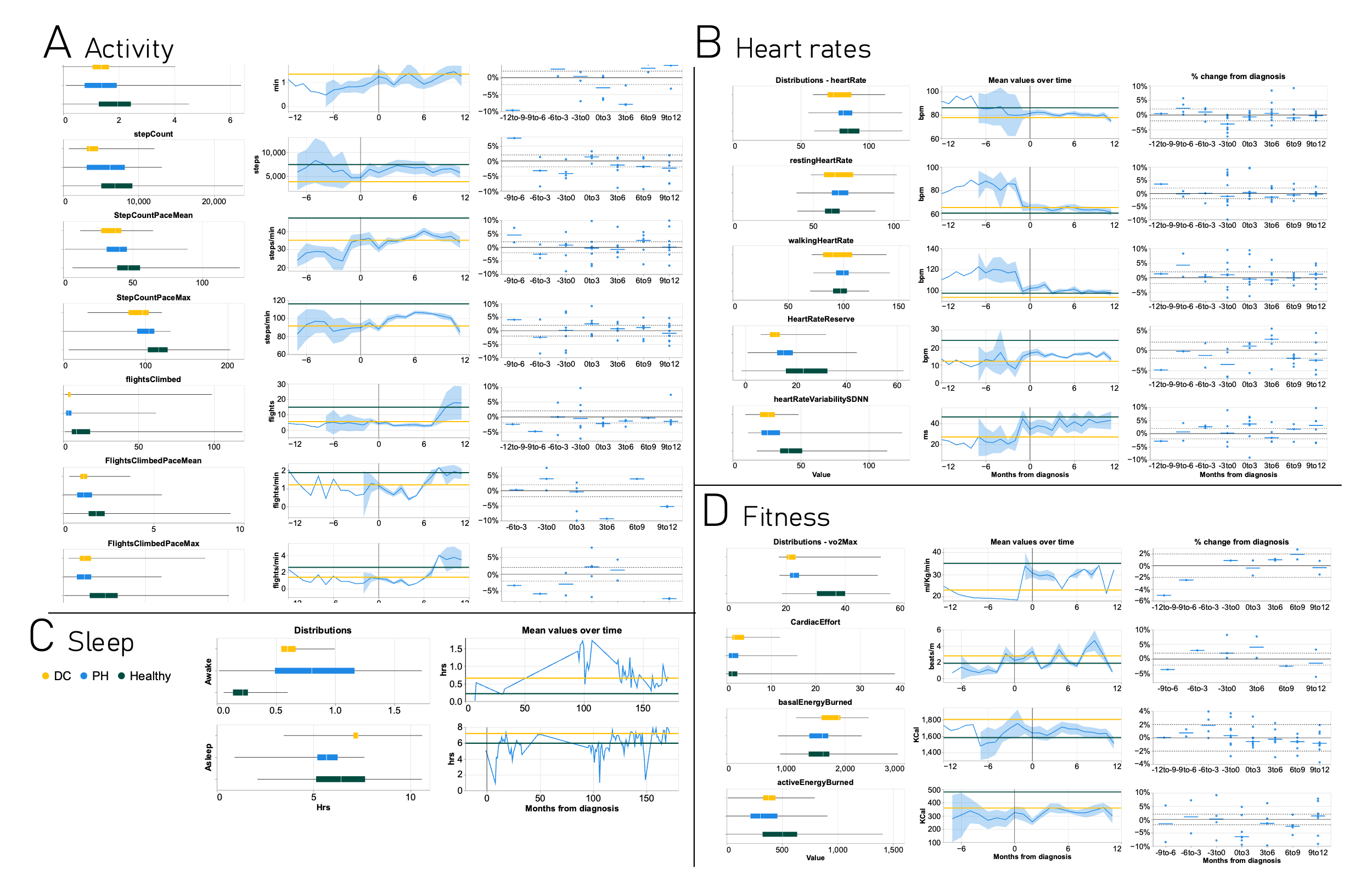

Figure SB2: description of Apple Watch metrics: distributions of values prior to diagnosis in PH case only with top 0.5% removed (left column), monthly mean values with 95% CI 12 months before and after diagnosis (right column, lines for DC and Healthy represent the average of the group in the whole period); and % change from diagnosis in 6 months intervals (right column, dots and strokes represent each patient and the median respectively). * pvalue from UMann-Whitney test. A one-way ANOVA test on the slopes between 12 and 6 month prior to diagnosis and 0-6 months after diagnosis shows 90% significance $.

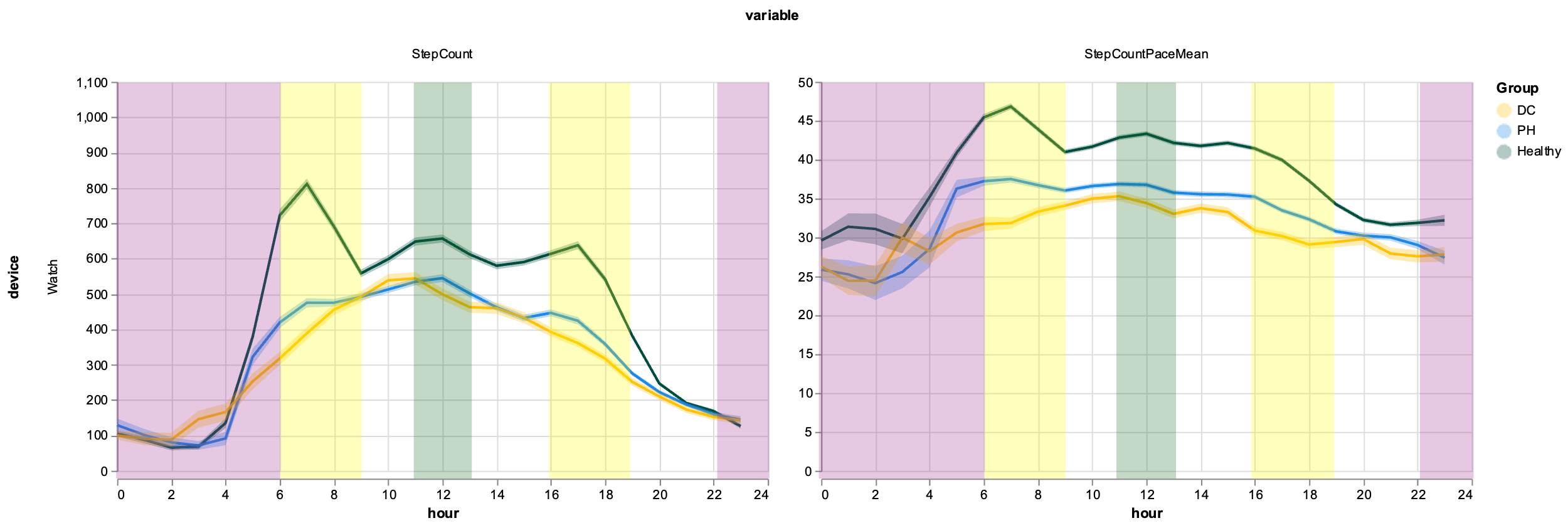

*Figure SB3: time of the day pooled distribution of Step Count and mean Gait Speed (Step Count Pace). Yellow bands are commute times, green is lunch time break, purple sleep time.*
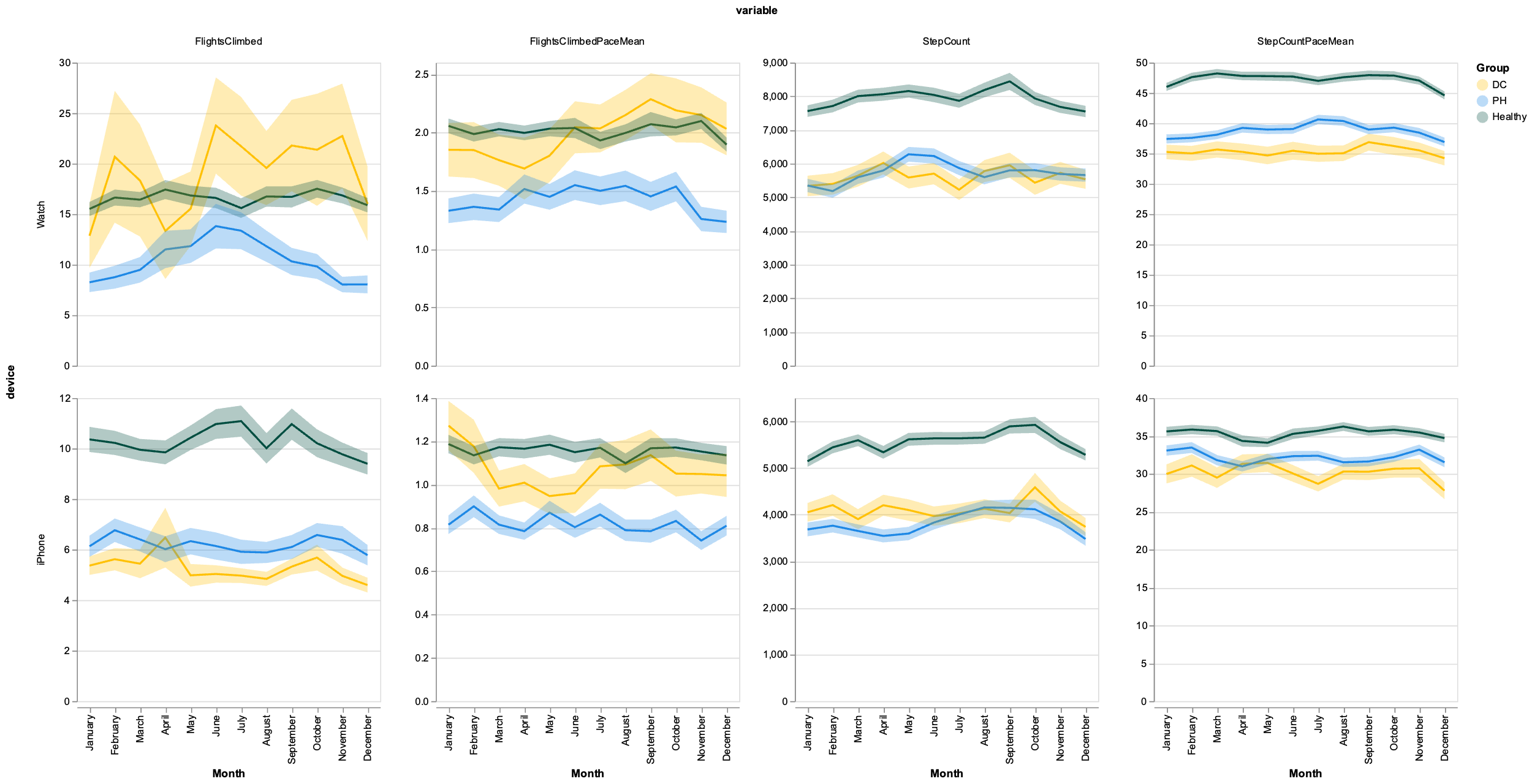

*Figure SB4: seasonality distribution for phone and watch Step Count and Flights Climbed metrics – pooled data*
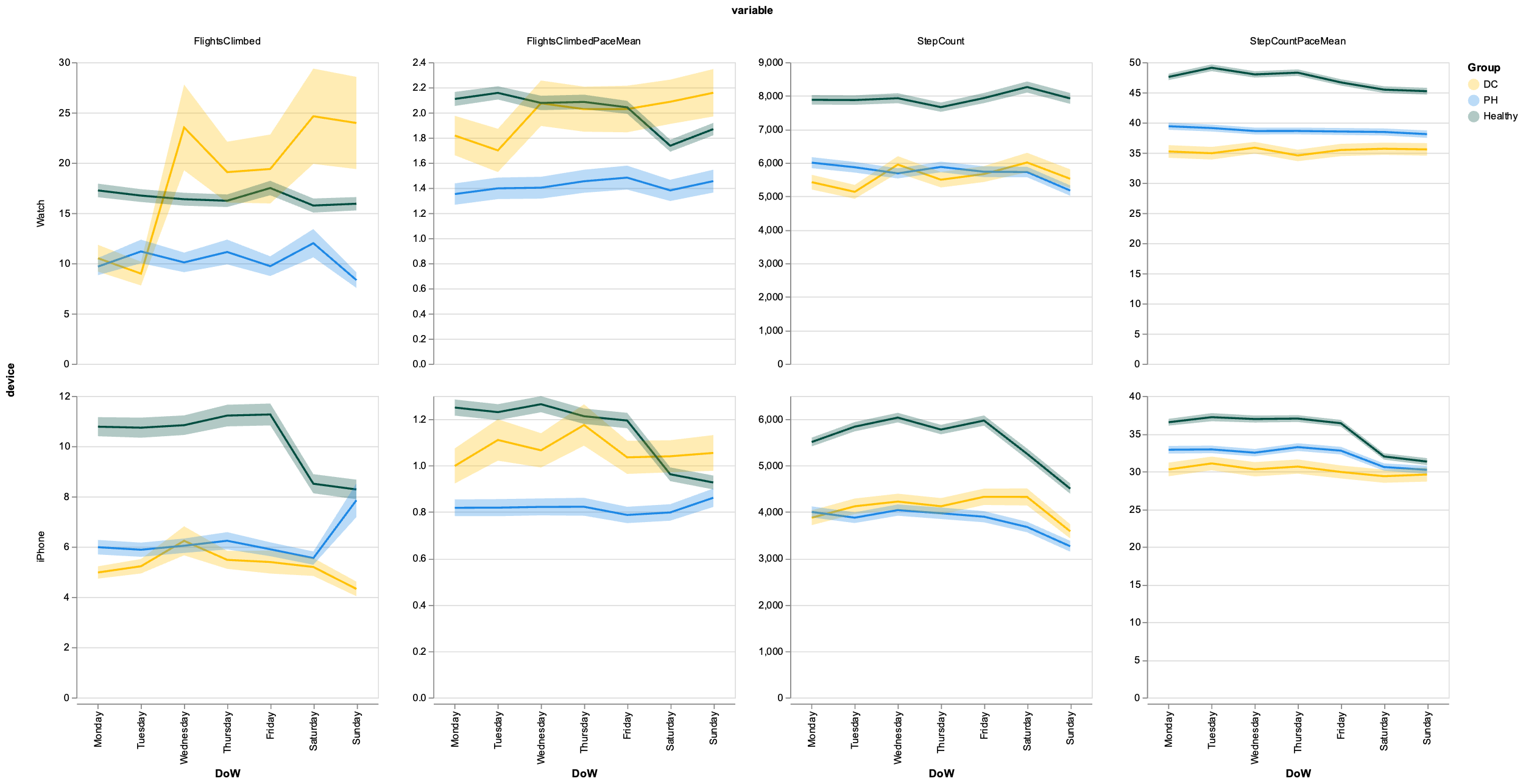

*Figure SB5: day of the week distribution for phone and watch Step Count and Flights Climbed metrics – pooled data*

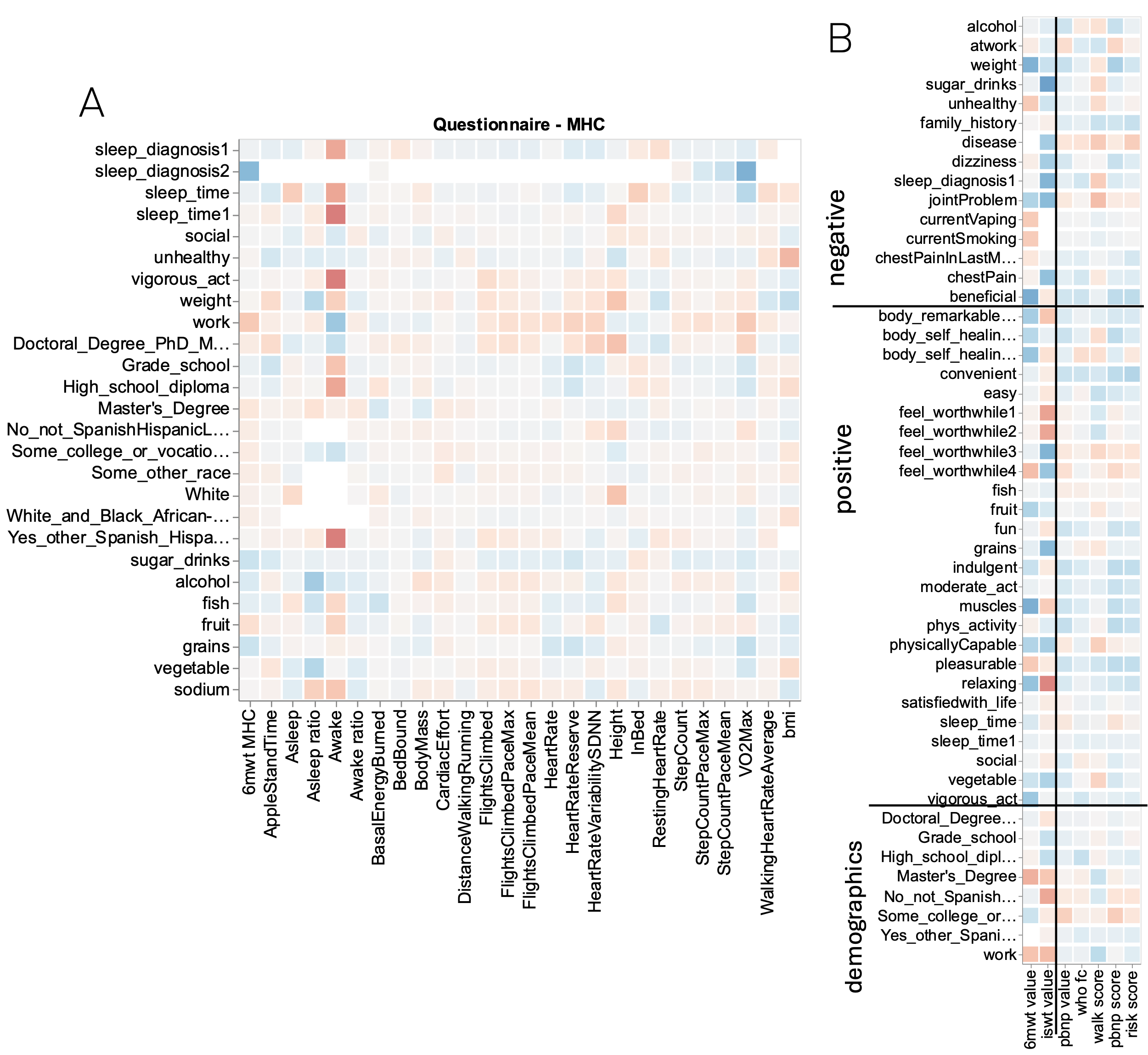

Figure SB6: Pearson’s correlations between variables: A) between watch and phone metrics and questionnaire answers; and B) between clinical walk tests + risk scores and questionnaire answers.

Appendix C: Model Meta-analysis

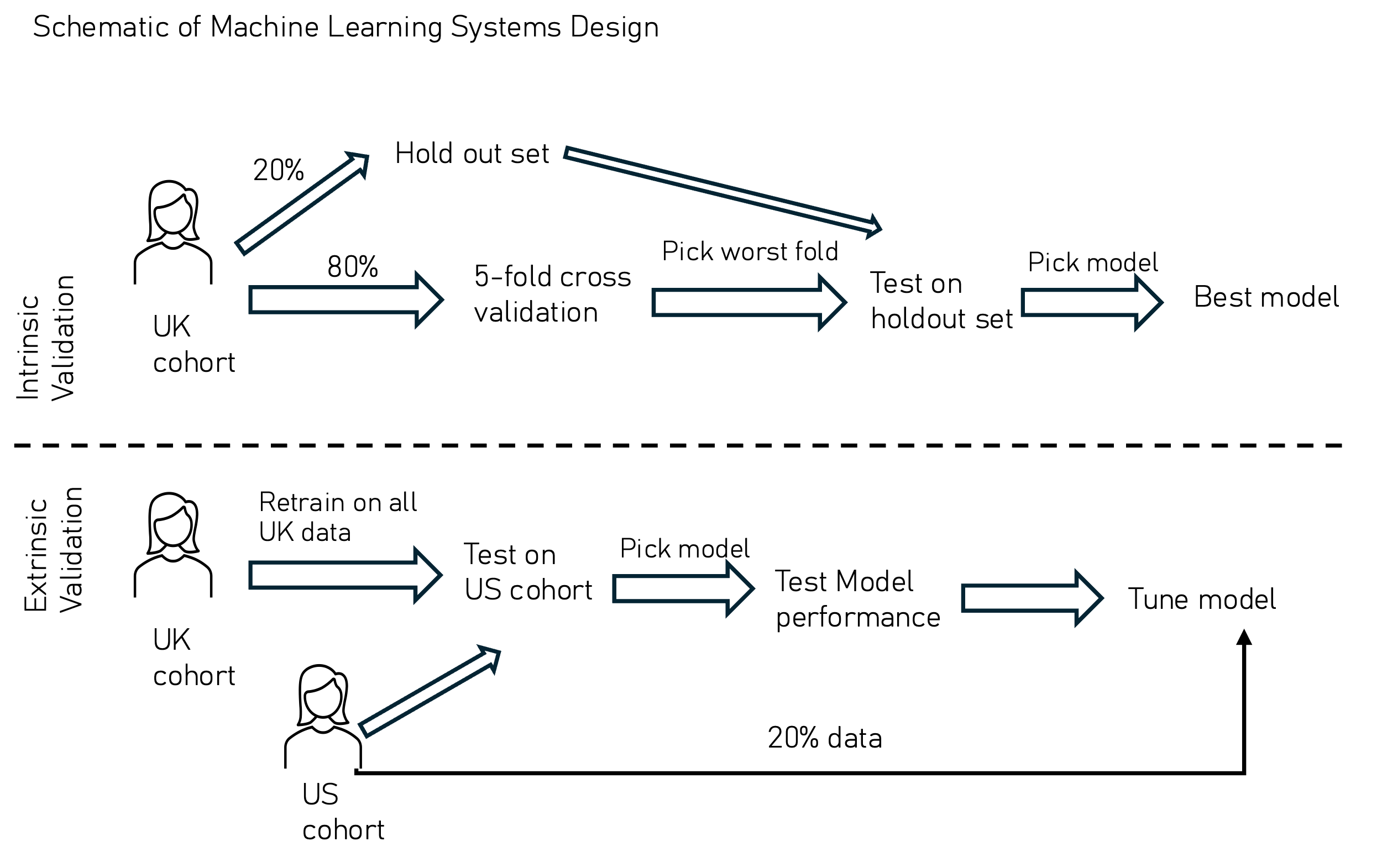

Figure SC1: schematic of validations

#### Comparison of Linear *SVM* vs XGBoost

Results of both linear SVM and XGBoost are sufficiently good, with ROC AUCs of 0.87 and 0.91 respectively in the example on Figure SC2. They are not overfitted as demonstrated by the narrow difference between train and test performance. F1 scores of thresholds chosen from the training set only tend to favour recall, which is preferred over Precision. However, a narrower difference between them might be desirable. Overall XGBoost provides better results than the linear SVM, hence why we chose it as the preferred method. It is also more resilient to missing values and data sparsity at the expense of being more prone to overfitting (which is not our case).

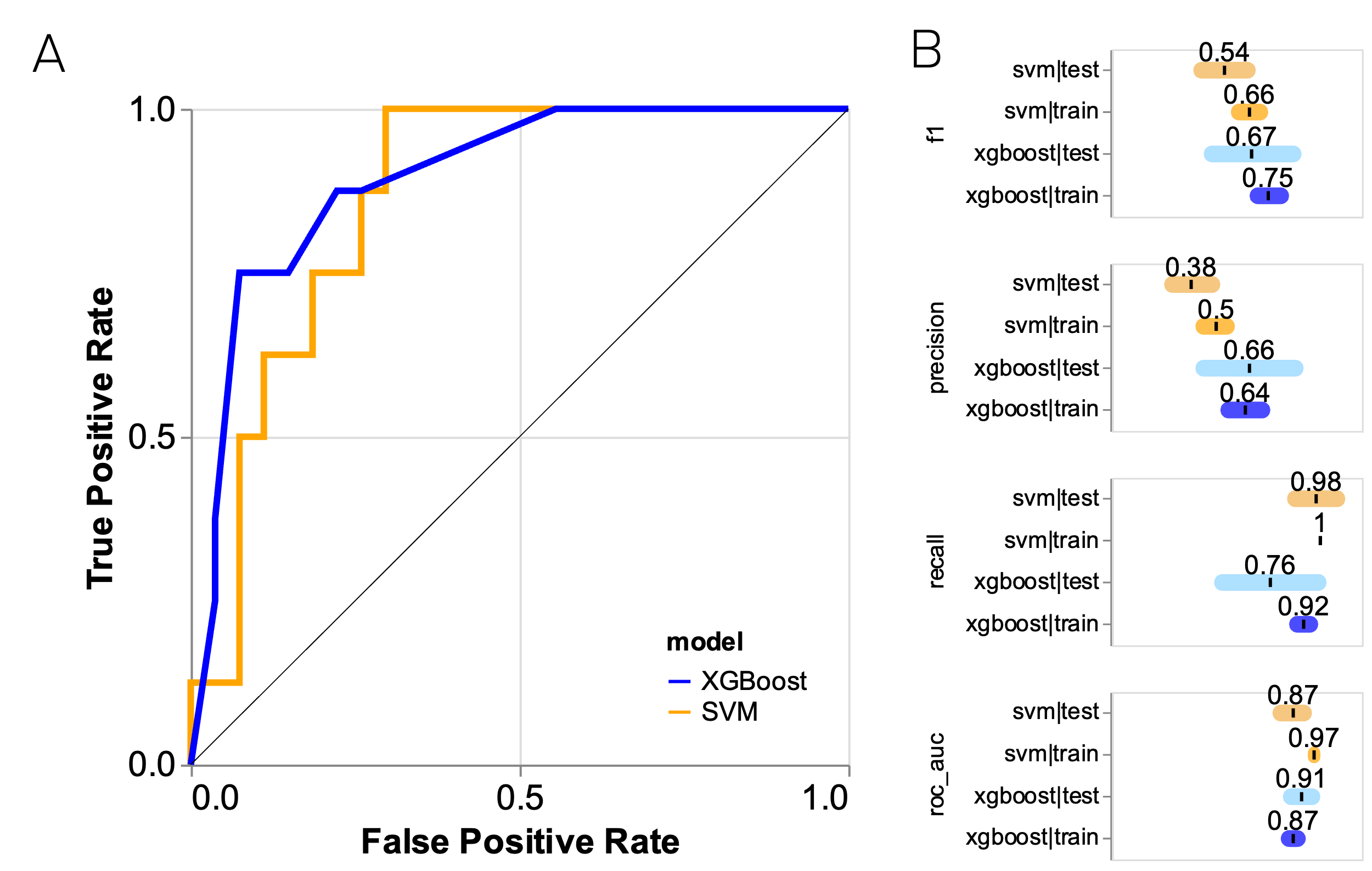

Figure SC2: comparison between models using phone + LifeStyle factors as an example feature set. A) ROC curve, B) scores represented as mean±std

#### US and UK dataset comparison

The drift in distribution was characterised by the Population Stability Index (PSI) all datasets except for the prediagnosis activity only show significant shift. Questionnaire metrics are the most variable.

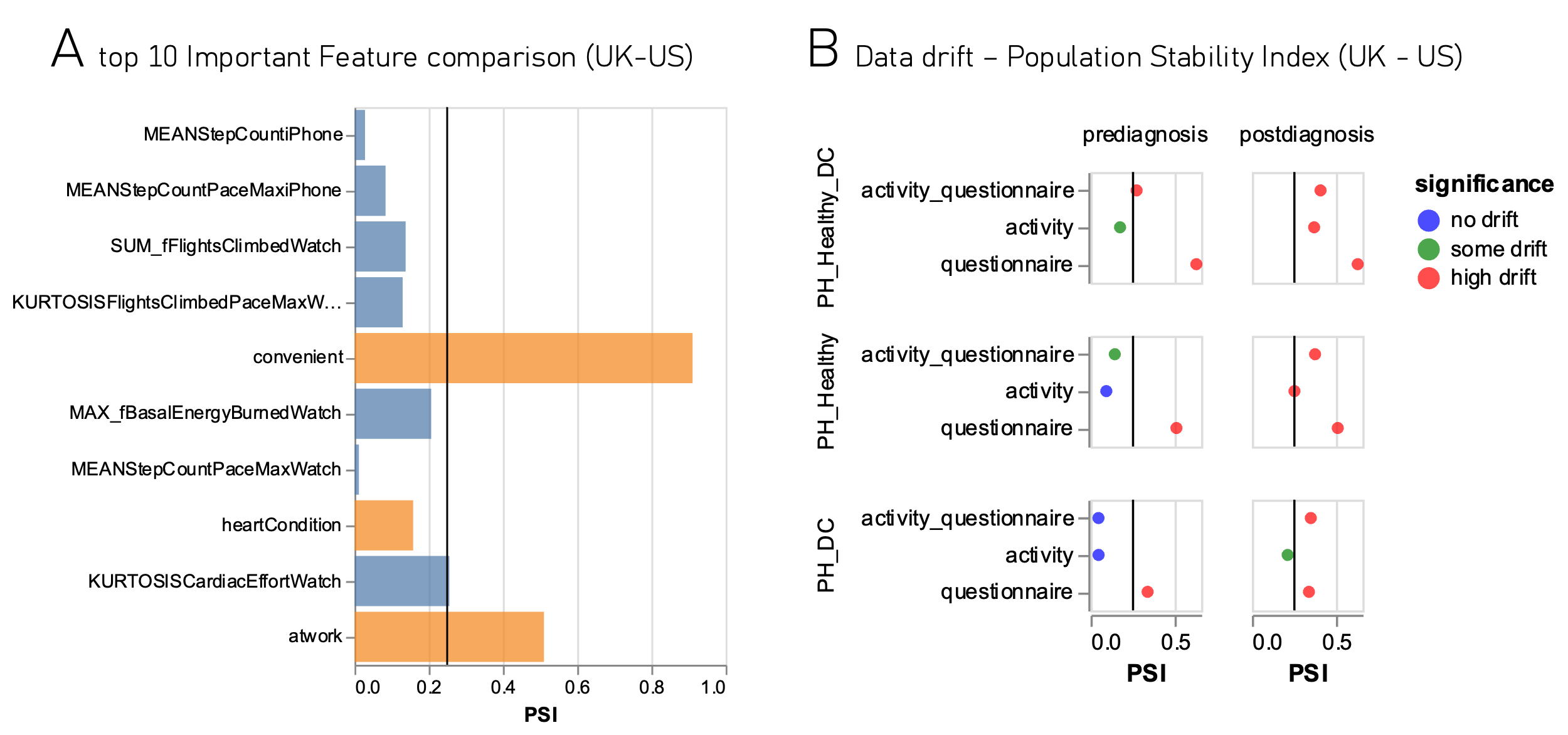

Figure SC3: Data drift by Population Stability Index – for top 10 metrics (A) and by dataset and optimization objective (B)

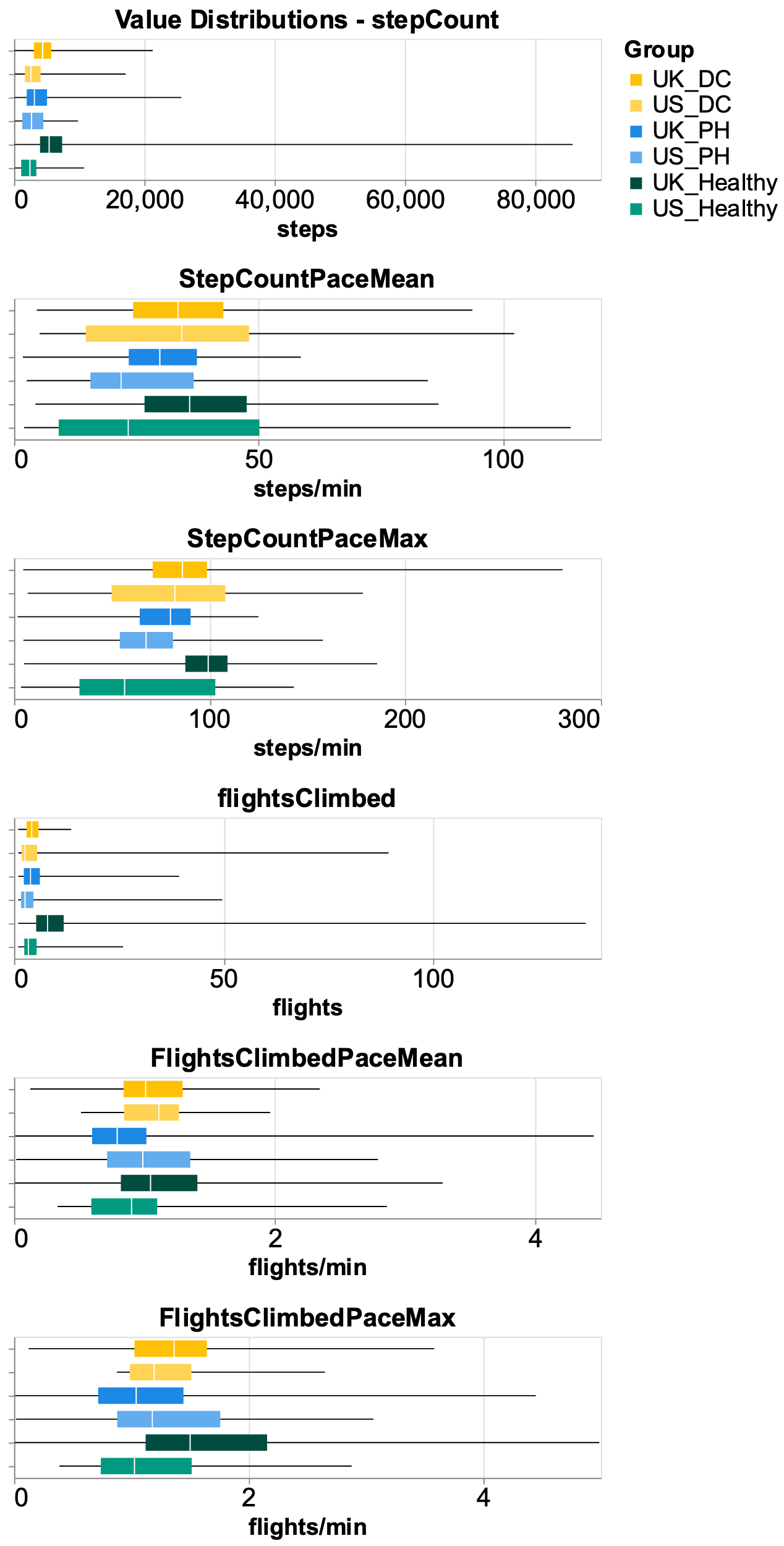

Figure SC4: comparison of UK and US distributions of iPhone metrics by metric and group.

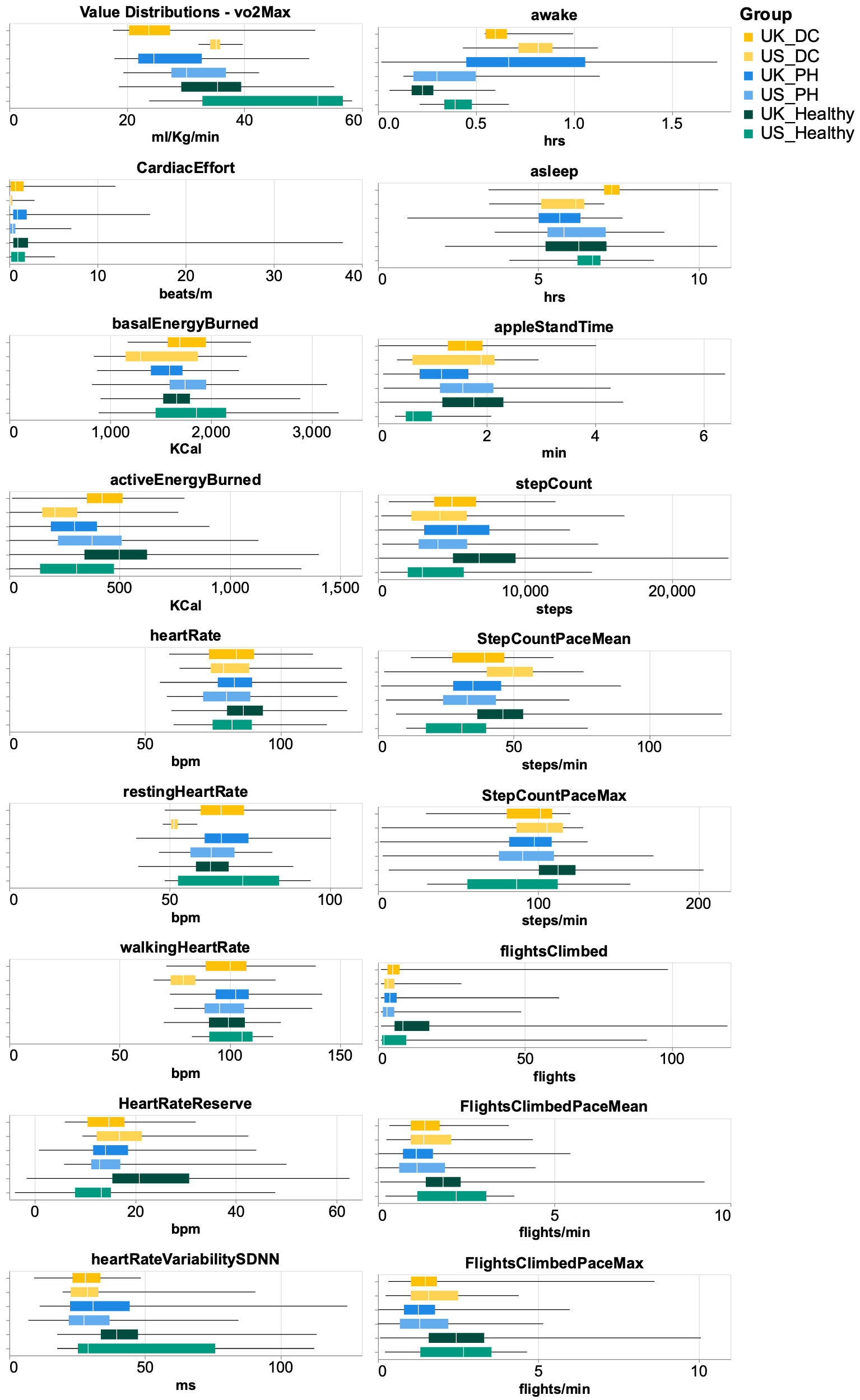

Figure SC5: comparison of UK and US distributions of watch metrics by metric and group.

#### Model retraining

The threshold of 20% was chosen as the minimum amount of data providing sufficient ROC AUC scores. This is dictated by the sharp increase from 10% to 20%.

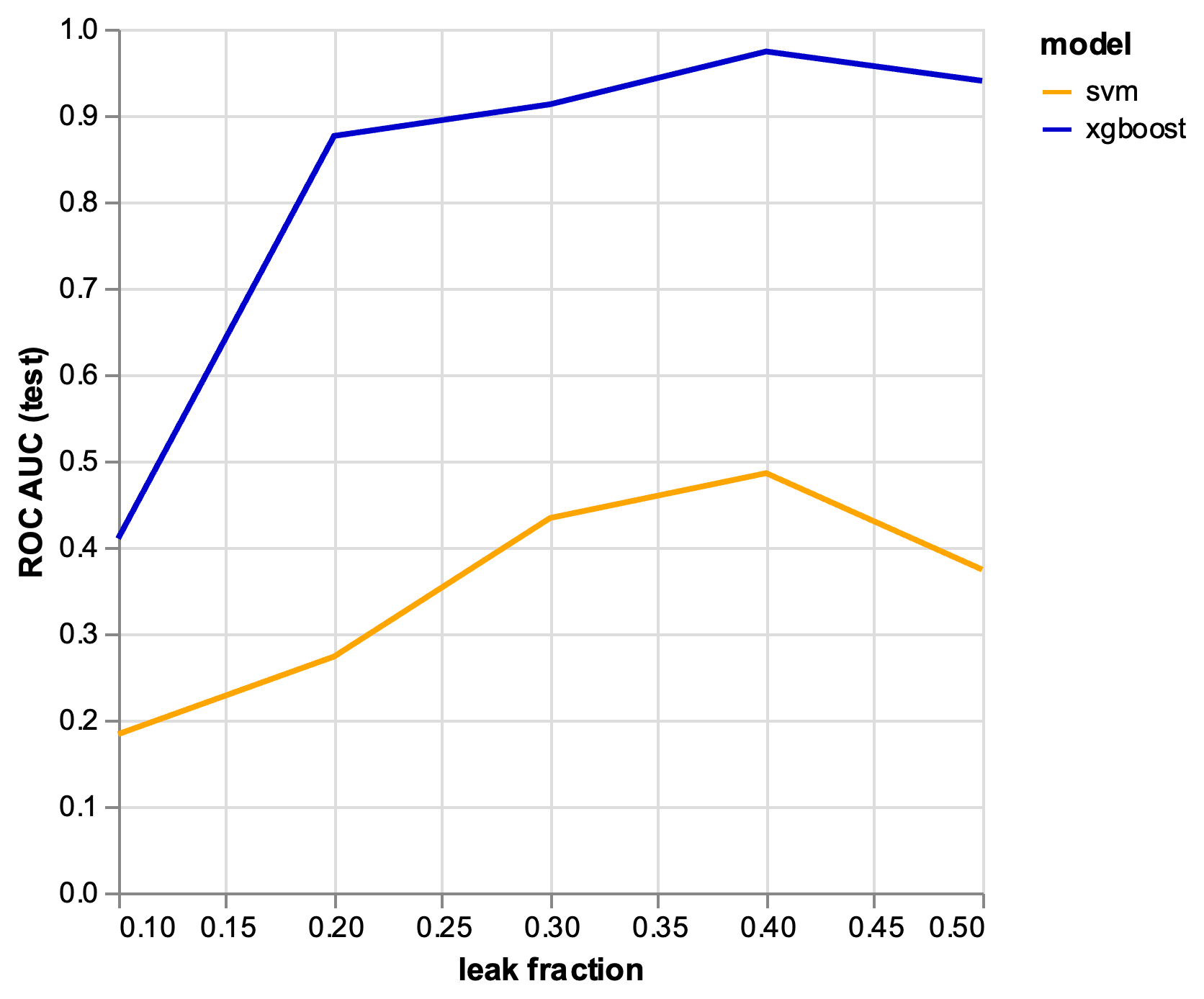

Figure SC7: ROC AUC for linear and xgboost model retraining at different ‘leak levels’

Table SC1: patient numbers for each activity feature set. * are not sufficient to fit the models.

|  | iPhone | Apple Watch |
| --- | --- | --- |
| All | | |
| Activity | 104 | 89 |
| Heart Rates | 0 | 92 |
| Fitness | 11 | 94 |
| Sleep | 0 | 34* |
| Prediagnosis | | |
| Activity | 91 | 70 |
| Heart Rates | 0 | 73 |
| Fitness | 3 | 75 |
| Sleep | 0 | 26* |

Table SC2: decision tree cumulative feature importance for final XGboost model.*metrics are the ones related to walking or climbing stairs.

| Features | Cumulative importance | Questionnaire Prompt |
| --- | --- | --- |
| MEANStepCountiPhone* | 0.17 |  |
| MEANStepCountPaceMaxiPhone* | 0.31 |  |
| SUM_fFlightsClimbedWatch* | 0.40 |  |
| KURTOSISFlightsClimbedPaceMaxWatch* | 0.48 |  |
| Convenient | 0.55 | EXERCISING is: convenient |
| MAX_fBasalEnergyBurnedWatch | 0.61 |  |
| MEANStepCountPaceMaxWatch* | 0.66 |  |
| heartCondition | 0.71 | has your doctor ever said that you have a heart condition and that you should only do physical activity recommended by a doctor? |
| KURTOSISCardiacEffortWatch | 0.75 |  |
| Atwork | 0.80 | Work Time Activity. |
| riskfactors2 | 0.83 | Over the next 10 years, compared to others your age and sex, how would you rate your risk of having a heart attack, stroke, or dying due to cardiovascular disease? (choose one) |
| currentSmoking | 0.86 | Do you currently smoke? |
| riskfactors1 | 0.88 | Over the next 10 years how likely do you think it is that you personally will have a heart attack, stroke, or die due to cardiovascular disease? (choose one) |
| CREST FACTORHeightiPhone | 0.90 |  |
| KURTOSISStepCountPaceMeaniPhone* | 0.91 |  |
| STDAppleStandTimeWatch | 0.92 |  |
| MINStepCountPaceMaxiPhone* | 0.93 |  |
| MAXFlightsClimbedPaceMaxWatch* | 0.94 |  |
| MINFlightsClimbediPhone* | 0.95 |  |
| graduate_school | 0.96 | What is the highest grade in school you finished? Choose the best answer. |
| sigma2StepCountPaceHigher70pctiPhone* | 0.97 |  |
| feel_worthwhile2 | 0.98 | How about happy? |
| MAXDistanceWalkingRunningiPhone* | 0.99 |  |
| CREST FACTORFlightsClimbedPaceMaxiPhone* | 1.00 |  |
| Fish | 1.00 | How many servings of fish do you eat on an average week? |
| MINActiveEnergyBurnedWatch | 1.00 |  |
| ar.L1FlightsClimbedWatch* | 1.00 |  |

### Appendix D: Questionnaire Meta-data

##### D1 Questionnaire Data Cleaning Pipeline

Questionnaire data was initially parsed and translated using…

Then the data was featurised and encoded following a mixture of numeric, ordinal, binary and translation rules.

**Numeric:** Variables like ‘pieces of fruit per day’, was left ‘as is’.

**Ordinal:** the variables with a clear ordinal sequence, distance between them are assumed to be equidistant, for instance:

{"Strongly Disagree":-3, "Disagree":-2, "Somewhat Disagree":-1, "Somewhat Agree":1, "Agree":2, "Strongly Agree":3}

Rules: The non-obvious ordinal answers were encoded based on judgement, as follows:

"atwork": {"I spent most of the day walking or using my hands and arms in work that required moderate exertion":2,

"I spent most of the day sitting or standing":0,

"I spent most of the day lifting or carrying heavy objects or moving most of my body in some other way":3},

"sodium":{"I avoid eating prepackaged and processed foods; and I avoid salt when I'm cooking at home":0,

'I avoid eating prepackaged and processed foods.':2,

'I avoid eating prepackaged and processed foods; and I avoid eating out, but when I do, I seek out low-sodium options':1,

"I avoid eating prepackaged and processed foods; and I avoid eating out, but when I do, I seek out low-sodium options; and I avoid salt when I'm cooking at home":0,

"I avoid salt when I'm cooking at home":1,

"I avoid eating out, but when I do, I seek out low-sodium options; and I avoid salt when I'm cooking at home":0,

'I avoid eating out, but when I do, I seek out low-sodium options.':1},

"beneficial": {"Moderately beneficial for my health":1,

"Very beneficial for my health":2,

"Slightly harmful for my health":-1,

"Moderately harmful for my health":-2,

"Neither harmful nor beneficial for my health":0,

"Extremely beneficial for my health":3,

"Very harmful for my health":-3},

"convenient": {"Somewhat convenient":1,

"Somewhat inconvenient":-1,

"Very convenient":2,

"Very inconvenient":-2},

"disease": {"Decreaess my risk slightly":1,

"Decreaess my risk moderately":2,

"Neither increases nor decreases my risk":0,

"Increases my risk moderately":-2,

"Increases my risk slightly":-1,

"Decreaess my risk very much":3,

"Increases my risk very much":-3},

"easy": {"Somewhat difficult":-1,

"Very easy":2,

"Very difficult":-1,

"Somewhat easy":1},

"fun": {"Somewhat fun":1,

"Very fun":2,

"Somewhat boring":-1,

"Very boring":-2},

"indulgent": {"Somewhat indulgent":1,

"Somewhat depriving":-1,

"Very indulgent":2,

"Very depriving":-2},

"muscles": {"Strengthening slightly":1,

"Strengthening moderately":2,

"Weakening slightly":-1,

"Neither strengthening nor weakening":0,

"Strengthening very much":3,

"Weakening moderately":-2,

"Weakening very much":-3},

"phys_activity": {"About three times a week, I did moderate activities":2,

"Once or twice a week, I did light activities":1,

"About three times a week, I did vigorous activities":3,

"I did not do much physical activity":0,

"Almost daily, that is five or more times a week, I did moderate activities":4,

"Almost daily, that is, five or more times a week, I did vigorous activities":5},

"pleasurable": {"Somewhat pleasurable":1,

"Very pleasurable":2,

"Very unpleasant":-2,

"Somewhat unpleasant":-1},

"relaxing": {"Somewhat relaxing":1,

"Very relaxing":2,

"Somewhat stressful":-1,

"Very stressful":-2},

"riskfactors1": {"A little":1,

"Moderately":2,

"Not at all":0,

"A lot":3,

"Extremely":4},

"riskfactors2": {"Higher than average":3,

"Much lower than average":0,

"Average":2,

"Lower than average":1,

"Much higher than average":4},

"riskfactors3": {"Moderately":2,

"A lot":3,

"A little":1,

"Extremely":4,

"Not at all":0},

"riskfactors4": {"Higher than average":3,

"Lower than average":1,

"Average":2,

"Much lower than average":0,

"Much higher than average":4},

"social": {"Somewhat social":1,

"Very lonely":-2,

"Somewhat lonely":-1,

"Very social":2}

}

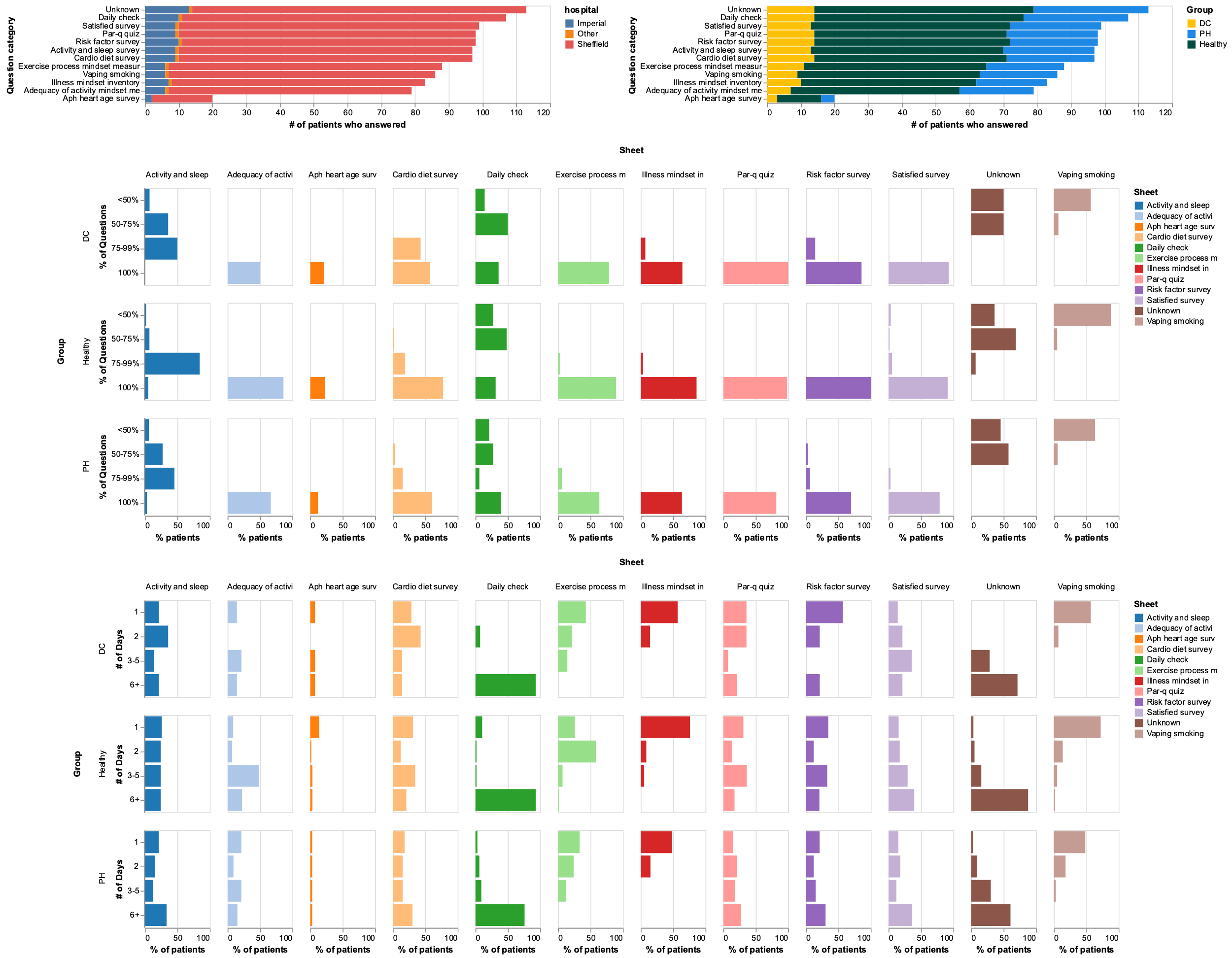

*Figure SD1: Distribution of questionnaire answers by Sheet*

*Table SD1: count of patients, questions and timepoints by questionnaire Sheet*

|  | **UK** | | | **US** | | |
| --- | --- | --- | --- | --- | --- | --- |
| **Sheet** | **patients** | **questions** | **timepoints** | **patients** | **questions** | **timepoints** |
| Vaping_Smoking | 88 | 12 | 37 | 21 | 10 | 25 |
| ACTIVITY AND SLEEP SURVEY | 112 | 9 | 1465 | 62 | 9 | 3451 |
| Illness_mindset_inventory | 85 | 20 | 34 | 20 | 20 | 20 |
| RISK FACTOR SURVEY | 101 | 7 | 205 | 93 | 7 | 175 |
| Exercise_process_mindset_measur | 90 | 7 | 61 | 22 | 7 | 37 |
| PAR-Q QUIZ | 100 | 7 | 205 | 137 | 7 | 175 |
| Adequacy_of_activity_mindset_me | 80 | 5 | 112 | 22 | 5 | 84 |
| DEMOGRAPHICS | 116 | 3 | 90 | 32 | 3 | 48 |
| CARDIO DIET SURVEY | 101 | 6 | 206 | 34 | 6 | 156 |
| DAILY CHECK | 111 | 10 | 1465 | 62 | 10 | 3451 |
| DAY ONE | 94 | 2 | 40 | 139 | 2 | 22 |
| SATISFIED SURVEY | 103 | 9 | 323 | 75 | 9 | 307 |
| APH HEART AGE SURVEY | 21 | 11 | 106 | 36 | 11 | 85 |

Table SD2: Top questions with major differences between PH, Healthy and DC groups for the UK cohort assessed by Kruskal-Wallis test. Last column indicates the variables that have been excluded from the machine learning model (*).

| **Question ID** | **Category** | **Prompt** | **Log10 pvalues** |
| --- | --- | --- | --- |
| **PAH*** | VASCULAR | Which vascular disease diagnosis have you received? | -16.6 |
| **Pulmonary_Hypertension*** | HEART | Have you been diagnosed with any of the below diseases? | -13.3 |
|  | DISEASE |  |  |
| **prescriptionDrugs*** | PAR-Q QUIZ | Is your doctor currently prescribing drugs (for example water pills) for your blood pressure or heart condition? | -12.7 |
| **medications_to_treat*** | RISK FACTOR SURVEY | Do you take medications to treat the following risk factors (indicate all that apply) | -9.2 |
| **heartCondition*** | PAR-Q QUIZ | has your doctor ever said that you have a heart condition and that you should only do physical activity recommended by a doctor? | -8.9 |
| **work** | ACTIVITY AND SLEEP SURVEY | Do you do regular work? | -6.2 |
| **riskfactors2** | SATISFIED SURVEY | Over the next 10 years, compared to others your age and sex, how would you rate your risk of having a heart attack, stroke, or dying due to cardiovascular disease? (choose one) | -5.8 |
| **riskfactors4** | SATISFIED SURVEY | Over your lifetime, compared to others your age and sex, how would you rate your risk of having a heart attack, stroke, or dying due to cardiovascular disease? (choose one) | -4.2 |
| **physicallyCapable** | PAR-Q QUIZ | Do you know of any reason why you should not do physical activity? | -4.1 |
| **riskfactors1** | SATISFIED SURVEY | Over the next 10 years how likely do you think it is that you personally will have a heart attack, stroke, or die due to cardiovascular disease? (choose one) | -4 |
| **riskfactors3** | SATISFIED SURVEY | Over your lifetime how likely do you think it is that you personally will have a heart attack, stroke, or die due to cardiovascular disease? (choose one) | -3.2 |
| **Stroke*** | VASCULAR | Which vascular disease diagnosis have you received? | -3.1 |
| **sugar_drinks** | CARDIO DIET SURVEY | How many beverages with added sugar do you drink every week? | -2.7 |
| **Grade_school** | education | What is the highest grade in school you finished? Choose the best answer. | -2.5 |
| **Heart_Failure_or_CHF_and_Pulmonary_Hypertension*** | HEART | Have you been diagnosed with any of the below diseases? | -2.1 |
|  | DISEASE |  |  |
| **chronic_illness_more_meaning_in_life** | Illness | Having a chronic illness allows you to find more meaning in life. | -2 |
|  | Mindset |  |  |
|  | inventory |  |  |
| **relaxing** | Exercise | EXERCISING is: relaxing | -2 |
|  | Process |  |  |
|  | Mindset |  |  |
| **Atrial_fibrillation_Afib*** | HEART | Have you been diagnosed with any of the below diseases? | -1.8 |
|  | DISEASE |  |  |
| **fruit** | CARDIO DIET SURVEY | How many cups of fruit do you eat in an average day? | -1.7 |
| **body_self_healing_from_most_conditions_and_diseases** | Illness_mindset_inventory | Your body is able to heal itself from most conditions and diseases. | -1.7 |
| **body_self_healing_in_many_different_circumstances** | Illness_mindset_inventory | Your body can heal itself on its own in many different circumstances. | -1.7 |
| **Atrial_fibrillation_Afib_and_Pulmonary_Hypertension*** | HEART | Have you been diagnosed with any of the below diseases? | -1.5 |
|  | DISEASE |  |  |
| **grains** | CARDIO DIET SURVEY | How many servings of whole grains do you eat on an average day? | -1.5 |
| **chronic_illness_handling** | Illness_mindset_inventory | A chronic illness is something that can be dealt with. | -1.4 |

The FAMD analysis shows visual separation between respondents.

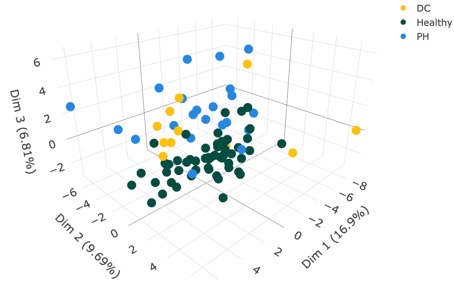

*Figure SD2: FAMD projections of questionnaire answers*

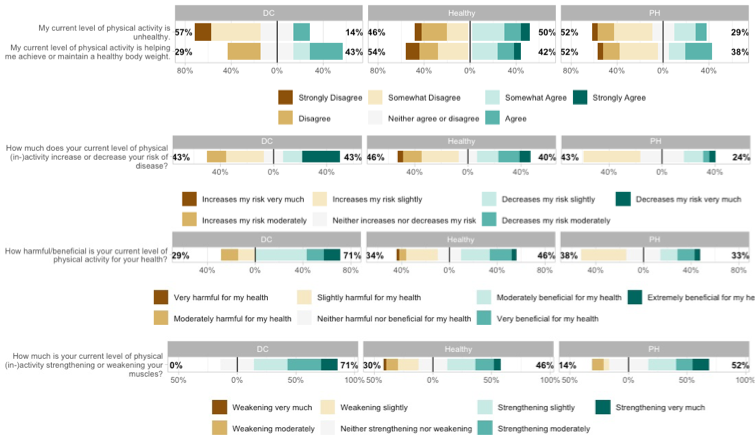

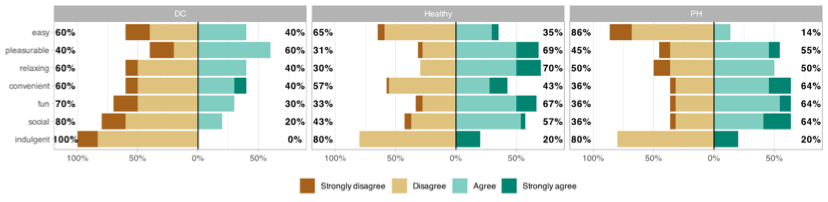

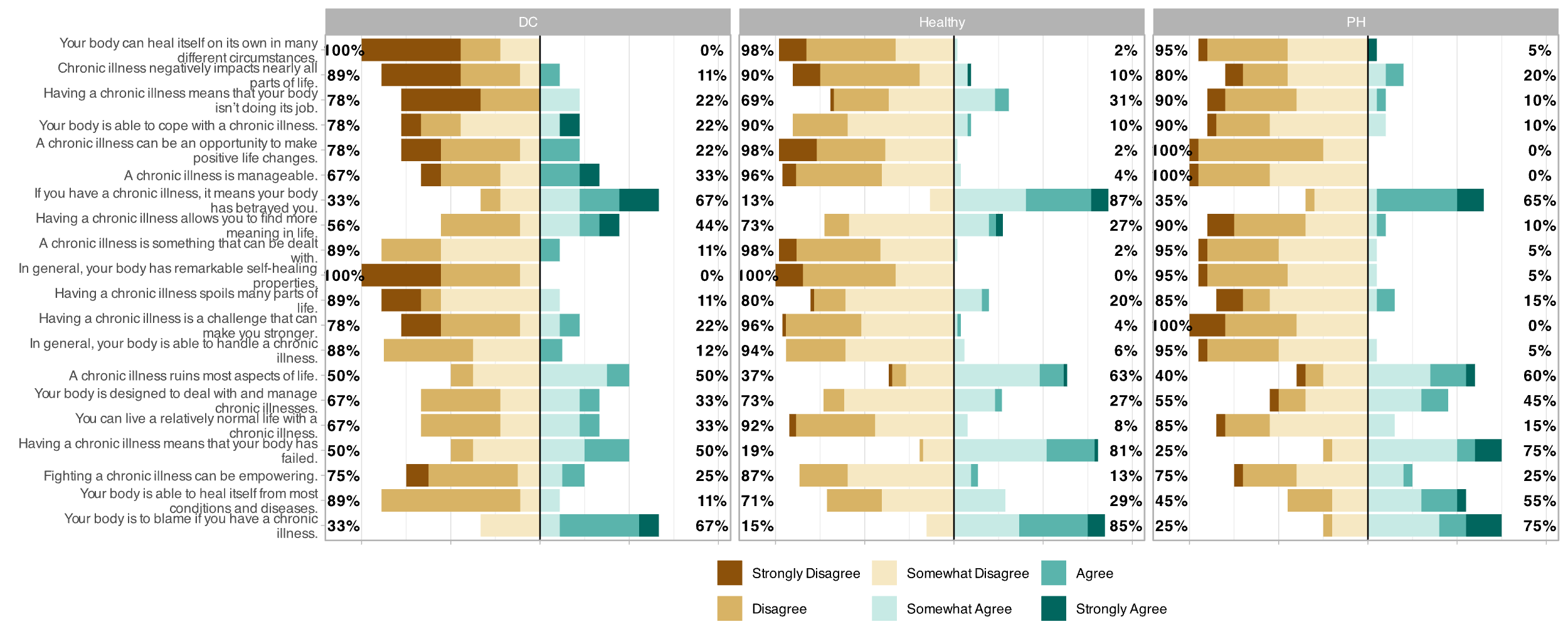

*Figure SD3: responses to questionnaires by group.*

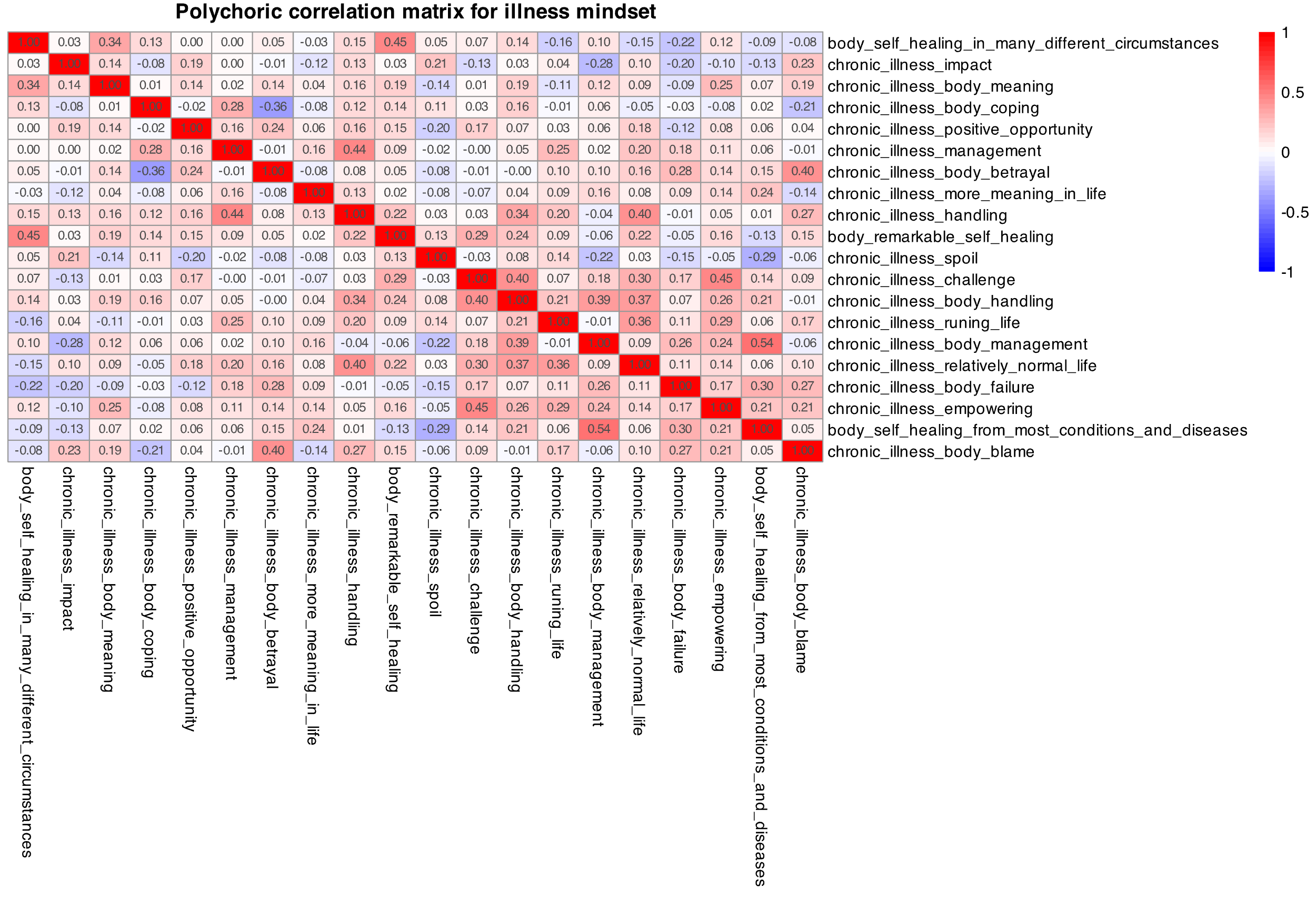

*Figure SD4: Polychoric correlation analysis of questionnaires.*

Table SD3: Questions in questionnaire with exclusions and groupings by Sheet (native to MHC) and used in modelling (Feature set)

| Feature set | Sheet | Question | exclude | Prompt |
| --- | --- | --- | --- | --- |
| PAR-Q QUIZ | PAR-Q QUIZ | chestPain | FALSE | Do you feel pain in your chest when you do physical activity? |
| PAR-Q QUIZ | PAR-Q QUIZ | chestPainInLastMonth | FALSE | In the past month, have you had chest pain when you were not doing physical activity? |
| PAR-Q QUIZ | PAR-Q QUIZ | dizziness | FALSE | Do you lose your balanced because of dizziness or do you ever lose consciousness? |
| PAR-Q QUIZ | PAR-Q QUIZ | heartCondition | FALSE | has your doctor ever said that you have a heart condition and that you should only do physical activity recommended by a doctor? |
| PAR-Q QUIZ | PAR-Q QUIZ | jointProblem | FALSE | Do you have a bone or joint problem that could be made worse by a change in your physical activity? |
| PAR-Q QUIZ | PAR-Q QUIZ | physicallyCapable | FALSE | Do you know of any reason why you should not do physical activity? |
| PAR-Q QUIZ | PAR-Q QUIZ | prescriptionDrugs | TRUE | Is your doctor currently prescribing drugs (for example water pills) for your blood pressure or heart condition? |
| LifeStyle | Vaping_Smoking | currentSmokeless | FALSE | Do you currently use smokeless tobacco (chewing tobacco, snuff, snus, and dissolvable tobacco products)? |
| LifeStyle | Vaping_Smoking | currentSmoking | FALSE | Do you current smoke cigarettes? |
| LifeStyle | Vaping_Smoking | currentVaping | FALSE | Do you currently vape nicotine (use ‚Äúe-cigs,‚Äù ‚Äúmods,‚Äù ‚Äúvape pens,‚Äù ‚Äúvapes‚Äù, ‚ÄúJUULs‚Äù, etc)? |
| LifeStyle | Vaping_Smoking | everQuitSmokeless | FALSE | During the past 12 months, have you tried to stop chewing tobacco (chewing tobacco, snuff, snus, and dissolvable tobacco products)?) |
| LifeStyle | Vaping_Smoking | everQuitSmoking | FALSE | During the past 12 months, have you tried to stop smoking cigarettes? |
| LifeStyle | Vaping_Smoking | everQuitVaping | FALSE | During the past 12 months, have you tried to stop vaping? |
| LifeStyle | Vaping_Smoking | onsetSmoking | FALSE | How old were you when you smoked your first tobacco cigarette (in years)? |
| LifeStyle | Vaping_Smoking | onsetVaping | FALSE | How old were you when you first vaped (in years)? |
| LifeStyle | Vaping_Smoking | pastSmokeless | FALSE | Have you used smokeless tobacco (chewing tobacco, snuff, snus, and dissolvable tobacco products) in the past? |
| LifeStyle | Vaping_Smoking | pastVaping | FALSE | Have you vaped in the past? |
| LifeStyle | Vaping_Smoking | readinessQuitSmokeless | FALSE | On a scale of 1-10, how ready do you feel to quit smokeless tobacco? |
| LifeStyle | Vaping_Smoking | readinessQuitSmoking | FALSE | On a scale of 1-10, how ready do you feel to quit smoking cigarettes? |
| LifeStyle | Vaping_Smoking | readinessQuitVaping | FALSE | On a scale of 1-10, how ready do you feel to quit vaping |
| LifeStyle | CARDIO DIET SURVEY | alcohol | FALSE | How many units of alcohol do you have per week? |
| LifeStyle | CARDIO DIET SURVEY | fish | FALSE | How many servings of fish do you eat on an average week? |
| LifeStyle | CARDIO DIET SURVEY | fruit | FALSE | How many cups of fruit do you eat in an average day? |
| LifeStyle | CARDIO DIET SURVEY | grains | FALSE | How many servings of whole grains do you eat on an average day? |
| LifeStyle | CARDIO DIET SURVEY | sugar_drinks | FALSE | How many beverages with added sugar do you drink every week? |
| LifeStyle | CARDIO DIET SURVEY | vegetable | FALSE | How many cups of vegetables do you eat in an average day? |
| LifeStyle | ACTIVITY AND SLEEP SURVEY | atwork | FALSE | Work Time Activity. |
| LifeStyle | ACTIVITY AND SLEEP SURVEY | moderate_act | FALSE | Overall, how many minutes of moderate activity do you get in a week? |
| LifeStyle | ACTIVITY AND SLEEP SURVEY | phys_activity | FALSE | Leisure Time Activity. |
| LifeStyle | ACTIVITY AND SLEEP SURVEY | sleep_diagnosis1 | FALSE | Have you ever been told by a doctor or other health professional that you have a sleep disorder? |
| LifeStyle | ACTIVITY AND SLEEP SURVEY | sleep_time | FALSE | How many hours of sleep did you get last night? |
| LifeStyle | ACTIVITY AND SLEEP SURVEY | sleep_time | FALSE | How much sleep do think you need every night to be rested? (in hours) |
| LifeStyle | ACTIVITY AND SLEEP SURVEY | sleep_time1 | FALSE | How much sleep do you usually get at night on weekdays or workdays? |
| LifeStyle | ACTIVITY AND SLEEP SURVEY | vigorous_act | FALSE | Overall, how many minutes of vigorous activity do you get in a week? |
| LifeStyle | ACTIVITY AND SLEEP SURVEY | work | FALSE | Do you do regular work? |
| Satisfied Survey | SATISFIED SURVEY | feel_worthwhile1 | FALSE | Overall, to what extent do you feel the things you do in your life are worthwhile? |
| Satisfied Survey | SATISFIED SURVEY | feel_worthwhile2 | FALSE | How about happy? |
| Satisfied Survey | SATISFIED SURVEY | feel_worthwhile3 | FALSE | How about worried? |
| Satisfied Survey | SATISFIED SURVEY | feel_worthwhile4 | FALSE | How about depressed? |
| Satisfied Survey | SATISFIED SURVEY | riskfactors1 | FALSE | Over the next 10 years how likely do you think it is that you personally will have a heart attack, stroke, or die due to cardiovascular disease? (choose one) |
| Satisfied Survey | SATISFIED SURVEY | riskfactors2 | FALSE | Over the next 10 years, compared to others your age and sex, how would you rate your risk of having a heart attack, stroke, or dying due to cardiovascular disease? (choose one) |
| Satisfied Survey | SATISFIED SURVEY | riskfactors3 | FALSE | Over your lifetime how likely do you think it is that you personally will have a heart attack, stroke, or die due to cardiovascular disease? (choose one) |
| Satisfied Survey | SATISFIED SURVEY | riskfactors4 | FALSE | Over your lifetime, compared to others your age and sex, how would you rate your risk of having a heart attack, stroke, or dying due to cardiovascular disease? (choose one) |
| Satisfied Survey | SATISFIED SURVEY | satisfiedwith_life | FALSE | Overall, how satisfied are you with life as a whole these days? |
| Activity Mindset | Illness_mindset_inventory | body_remarkable_self_healing | FALSE | In general, your body has remarkable self-healing properties. |
| Activity Mindset | Illness_mindset_inventory | body_self_healing_from_most_conditions_and_diseases | FALSE | Your body is able to heal itself from most conditions and diseases. |
| Activity Mindset | Illness_mindset_inventory | body_self_healing_in_many_different_circumstances | FALSE | Your body can heal itself on its own in many different circumstances. |
| Activity Mindset | Illness_mindset_inventory | chronic_illness_body_betrayal | FALSE | If you have a chronic illness, it means your body has betrayed you. |
| Activity Mindset | Illness_mindset_inventory | chronic_illness_body_blame | FALSE | Your body is to blame if you have a chronic illness. |
| Activity Mindset | Illness_mindset_inventory | chronic_illness_body_coping | FALSE | Your body is able to cope with a chronic illness. |
| Activity Mindset | Illness_mindset_inventory | chronic_illness_body_failure | FALSE | Having a chronic illness means that your body has failed. |
| Activity Mindset | Illness_mindset_inventory | chronic_illness_body_handling | FALSE | In general, your body is able to handle a chronic illness. |
| Activity Mindset | Illness_mindset_inventory | chronic_illness_body_management | FALSE | Your body is designed to deal with and manage chronic illnesses. |
| Activity Mindset | Illness_mindset_inventory | chronic_illness_body_meaning | FALSE | Having a chronic illness means that your body isn‚Äôt doing its job. |
| Activity Mindset | Illness_mindset_inventory | chronic_illness_challenge | FALSE | Having a chronic illness is a challenge that can make you stronger. |
| Activity Mindset | Illness_mindset_inventory | chronic_illness_empowering | FALSE | Fighting a chronic illness can be empowering. |
| Activity Mindset | Illness_mindset_inventory | chronic_illness_handling | FALSE | A chronic illness is something that can be dealt with. |
| Activity Mindset | Illness_mindset_inventory | chronic_illness_impact | FALSE | Chronic illness negatively impacts nearly all parts of life. |
| Activity Mindset | Illness_mindset_inventory | chronic_illness_management | FALSE | A chronic illness is manageable. |
| Activity Mindset | Illness_mindset_inventory | chronic_illness_more_meaning_in_life | FALSE | Having a chronic illness allows you to find more meaning in life. |
| Activity Mindset | Illness_mindset_inventory | chronic_illness_positive_opportunity | FALSE | A chronic illness can be an opportunity to make positive life changes. |
| Activity Mindset | Illness_mindset_inventory | chronic_illness_relatively_normal_life | FALSE | You can live a relatively normal life with a chronic illness. |
| Activity Mindset | Illness_mindset_inventory | chronic_illness_runing_life | FALSE | A chronic illness ruins most aspects of life. |
| Activity Mindset | Illness_mindset_inventory | chronic_illness_spoil | FALSE | Having a chronic illness spoils many parts of life. |
| Activity Mindset | Adequacy_of_activity_mindset_me | beneficial | FALSE | How harmful/beneficial is your current level of physical activity for your health? |
| Activity Mindset | Adequacy_of_activity_mindset_me | disease | FALSE | How much does your current level of physical (in-)activity increase or decrease your risk of disease? |
| Activity Mindset | Adequacy_of_activity_mindset_me | muscles | FALSE | How much is your current level of physical (in-)activity strengthening or weakening your muscles? |
| Activity Mindset | Adequacy_of_activity_mindset_me | unhealthy | FALSE | My current level of physical activity is unhealthy. |
| Activity Mindset | Adequacy_of_activity_mindset_me | weight | FALSE | My current level of physical activity is helping me achieve or maintain a healthy body weight. |
| Activity Mindset | Exercise_process_mindset_measur | convenient | FALSE | EXERCISING is: convenient |
| Activity Mindset | Exercise_process_mindset_measur | easy | FALSE | EXERCISING is: easy |
| Activity Mindset | Exercise_process_mindset_measur | fun | FALSE | EXERCISING is: fun |
| Activity Mindset | Exercise_process_mindset_measur | indulgent | FALSE | EXERCISING is: indulgent |
| Activity Mindset | Exercise_process_mindset_measur | pleasurable | FALSE | EXERCISING is: pleasurable |
| Activity Mindset | Exercise_process_mindset_measur | relaxing | FALSE | EXERCISING is: relaxing |
| Activity Mindset | Exercise_process_mindset_measur | social | FALSE | EXERCISING is: social |
| Risk factors | RISK FACTOR SURVEY | family_history | FALSE | Do you have a family history of early heart disease? |
| Risk factors | RISK FACTOR SURVEY | medications_to_treat | TRUE | Do you take medications to treat the following risk factors (indicate all that apply) |
| Risk factors | RISK FACTOR SURVEY | education_College_graduate_or_Baccalaureate_Degree | FALSE |  |
| Risk factors | RISK FACTOR SURVEY | education_Doctoral_Degree_PhD_MD_JD_etc | FALSE |  |
| Risk factors | RISK FACTOR SURVEY | education_Grade_school | FALSE |  |
| Risk factors | RISK FACTOR SURVEY | education_High_school_diploma | FALSE |  |
| Risk factors | RISK FACTOR SURVEY | education_Master's_Degree | FALSE |  |
| Risk factors | RISK FACTOR SURVEY | education_Some_college_or_vocational_school_or_Associate_Degree | FALSE |  |
| Risk factors | RISK FACTOR SURVEY | ethnicity_No_not_SpanishHispanicLatino | FALSE |  |
| Risk factors | RISK FACTOR SURVEY | ethnicity_Yes_Cuban | FALSE |  |
| Risk factors | RISK FACTOR SURVEY | ethnicity_Yes_other_Spanish_Hispanic_Latina | FALSE |  |
| Risk factors | RISK FACTOR SURVEY | heart_disease_Angina_heart_chest_pains | TRUE |  |
| Risk factors | RISK FACTOR SURVEY | heart_disease_Atrial_fibrillation_Afib | TRUE |  |
| Risk factors | RISK FACTOR SURVEY | heart_disease_Congenital_Heart | TRUE |  |
| Risk factors | RISK FACTOR SURVEY | heart_disease_Coronary_BlockageStenosis | TRUE |  |
| Risk factors | RISK FACTOR SURVEY | heart_disease_Coronary_StentAngioplasty | TRUE |  |
| Risk factors | RISK FACTOR SURVEY | heart_disease_Heart_AttackMyocardial_Infarction | TRUE |  |
| Risk factors | RISK FACTOR SURVEY | heart_disease_Heart_Bypass_Surgery | TRUE |  |
| Risk factors | RISK FACTOR SURVEY | heart_disease_Heart_Failure_or_CHF | TRUE |  |
| Risk factors | RISK FACTOR SURVEY | heart_disease_High_Coronary_Calcium_Score | TRUE |  |
| Risk factors | RISK FACTOR SURVEY | heart_disease_None_of_the_above | TRUE |  |
| Risk factors | RISK FACTOR SURVEY | heart_disease_Pulmonary_Hypertension | TRUE |  |
| Risk factors | RISK FACTOR SURVEY | heart_disease_nan | TRUE |  |
| Risk factors | RISK FACTOR SURVEY | race_Asian_Indian | FALSE |  |
| Risk factors | RISK FACTOR SURVEY | race_Black_African-American_or_Negro | FALSE |  |
| Risk factors | RISK FACTOR SURVEY | race_Chinise | FALSE |  |
| Risk factors | RISK FACTOR SURVEY | race_Filipino | FALSE |  |
| Risk factors | RISK FACTOR SURVEY | race_Some_other_race | FALSE |  |
| Risk factors | RISK FACTOR SURVEY | race_White | FALSE |  |
| Risk factors | RISK FACTOR SURVEY | race_White_and_Black_African-American_or_Negro_and_American_Indian | FALSE |  |
| Risk factors | RISK FACTOR SURVEY | race_White_and_Pacific_Islander | FALSE |  |
| Risk factors | RISK FACTOR SURVEY | vascular_PAH | TRUE |  |
| Risk factors | RISK FACTOR SURVEY | vascular_Abdominal_Aortic_Aneurysm | TRUE |  |
| Risk factors | RISK FACTOR SURVEY | vascular_Carotid_Artery_BlockageStenosis | TRUE |  |
| Risk factors | RISK FACTOR SURVEY | vascular_Carotid_Artery_Surgery_or_Stent | TRUE |  |
| Risk factors | RISK FACTOR SURVEY | vascular_None_of_the_above | TRUE |  |
| Risk factors | RISK FACTOR SURVEY | vascular_Peripheral_Vascular_Disease_BlockageStenosis_Surgery_or_Stent | TRUE |  |
| Risk factors | RISK FACTOR SURVEY | vascular_Stroke | TRUE |  |
| Risk factors | RISK FACTOR SURVEY | vascular_Transient_Ischemic_Attack_TIA | TRUE |  |
| Risk factors | RISK FACTOR SURVEY | vascular_nan | TRUE |  |
| Risk factors | DAILY CHECK | sleep_time | FALSE | How many hours of sleep did you get last night? |
| Risk factors | DAILY CHECK | sleep_time | FALSE | How much sleep do think you need every night to be rested? (in hours) |
| Risk factors | DEMOGRAPHICS | countryCode_CA | FALSE |  |
| Risk factors | DEMOGRAPHICS | countryCode_DE | FALSE |  |
| Risk factors | DEMOGRAPHICS | countryCode_GB | FALSE |  |
| Risk factors | DEMOGRAPHICS | countryCode_SE | FALSE |  |
| Risk factors | DEMOGRAPHICS | countryCode_SG | FALSE |  |
| Risk factors | DEMOGRAPHICS | countryCode_US | FALSE |  |
| Risk factors | DEMOGRAPHICS | patientFitzpatrickSkinType_Type_I | FALSE |  |
| Risk factors | DEMOGRAPHICS | patientFitzpatrickSkinType_Type_II | FALSE |  |
| Risk factors | DEMOGRAPHICS | patientBloodType_A+ | FALSE |  |
| Risk factors | DEMOGRAPHICS | patientBloodType_A- | FALSE |  |
| Risk factors | DEMOGRAPHICS | patientBloodType_O+ | FALSE |  |
| Risk factors | DEMOGRAPHICS | patientBloodType_O- | FALSE |  |

### Appendix E: Environment

altair==5.3.0

attrs==23.2.0

cachetools==5.3.3

certifi==2024.6.2

charset-normalizer==3.3.2

contourpy==1.2.1

cycler==0.12.1

db-dtypes==1.2.0

fonttools==4.53.0

google-api-core==2.19.0

google-auth==2.30.0

google-auth-oauthlib==1.2.0

google-cloud-bigquery==3.24.0

google-cloud-core==2.4.1

google-crc32c==1.5.0

google-resumable-media==2.7.1

googleapis-common-protos==1.63.1

grpcio==1.64.1

grpcio-status==1.62.2

idna==3.7

Jinja2==3.1.4

joblib==1.4.2

jsonschema==4.22.0

jsonschema-specifications==2023.12.1

kiwisolver==1.4.5

MarkupSafe==2.1.5

matplotlib==3.9.0

numpy==1.26.4

oauthlib==3.2.2

packaging==24.1

pandas==2.2.1

pandas-gbq==0.23.1

patsy==0.5.6

pillow==10.3.0

proto-plus==1.23.0

protobuf==4.25.3

pyarrow==16.1.0

pyasn1==0.6.0

pyasn1_modules==0.4.0

pydata-google-auth==1.8.2

pyparsing==3.1.2

python-dateutil==2.8.2

pytz==2024.1

referencing==0.35.1

requests==2.32.3

requests-oauthlib==2.0.0

rpds-py==0.18.1

rsa==4.9

scikit-learn==1.5.0

scipy==1.13.1

six==1.16.0

statsmodels==0.14.2

threadpoolctl==3.5.0

toolz==0.12.1

tqdm==4.66.2

tzdata==2024.1

urllib3==2.2.1

xgboost==2.0.3
